# Activated Dendritic Cell Subsets Characterize Muscle of Inclusion Body Myositis Patients and Correlate with KLRG1+ and TBX21+ CD8+ T cells

**DOI:** 10.1101/2025.06.04.25328910

**Authors:** Raphael A. Kirou, Iago Pinal-Fernandez, Maria Casal-Dominguez, Katherine Pak, Chiseko Ikenaga, Christopher Nelke, Sven Wischnewski, Stefania Del Orso, Faiza Naz, Shamima Islam, Gustavo Gutierrez-Cruz, Thomas E. Lloyd, Lucas Schirmer, Tobias Ruck, Werner Stenzel, Albert Selva-O’Callaghan, Jose C. Milisenda, Andrew L. Mammen

## Abstract

Inclusion body myositis (IBM) is an idiopathic inflammatory myopathy characterized by muscle-infiltrating KLRG1+ and TBX21+ cytotoxic T cells and type 1 inflammation. Myeloid dendritic cells (mDCs), including type 1 conventional dendritic (cDC1) cells, type 2 conventional dendritic (cDC2) cells, and mature immunoregulatory dendritic (mregDC) cells, have previously been reported in skeletal muscle of IBM patients and may activate these cytotoxic T cells. Here, we analyzed single-nucleus RNA-sequencing (snRNA-seq) and bulk RNA-sequencing (RNA-seq) data from skeletal muscle of IBM, other myositis, and control patients to identify and quantify these mDC subsets and characterize their contribution to IBM inflammation. Our findings reveal that all three mDC subsets are relatively increased and activated in muscle of IBM patients and correlate with IBM-specific inflammatory markers. Our data specifically implicates cDC1 cells in CD8+ T cell activation via specific expression of both KLRG1 ligands, *CDH1* and *CDH2*, as well as *IL12B* in IBM muscle.

## INTRODUCTION

Inclusion body myositis (IBM) is one of a large collection of inflammatory myopathies, which also includes dermatomyositis (DM), immune-mediated necrotizing myopathy (IMNM), anti-synthetase syndrome (ASyS), and overlap myositis, as well as polymyositis (PM), now considered a diagnosis of exclusion^1^. Although various immunomodulating agents have shown effectiveness for the other inflammatory myopathies, none have done so for IBM^2^. This has led to a multitude of studies aimed at understanding the pathophysiology of IBM and its immune-mediated muscle damage, in order to identify treatment targets. A common theme that has emerged is an overactive type 1 inflammatory response, with increased expression of Type II Interferon (IFN-II, or *IFNG*) and IFN-II-inducible genes^3, 4, 5, 6, 7, 8, 9, 10^. This seems to be mediated in large part by highly differentiated CD8+ T cells, including those expressing the immune checkpoint *KLRG1* and those expressing *TBX21*, the defining transcription factor of type 1 inflammation^5, 7, 11, 12^. However, it remains unclear how and why these cells are activated.

A likely source of CD8+ T cell activation are myeloid dendritic cells (mDCs), a group of professional antigen-presenting cells. In recent years, there has been an explosion of high-throughput sequencing studies describing subsets of mDCs. The first widely accepted subset is the type 1 conventional dendritic (cDC1) cells that are said to activate CD8+ T cell subsets and express markers including *CADM1*, *CLEC9A*, and *XCR1*^13, 14, 15, 16^. The second subset, the type 2 conventional dendritic (cDC2) cells, are characterized by expression of *CD1C*, and are said to activate CD4+ T cells^13, 14, 15, 16^. Numerous studies have discovered heterogeneity within CD1C+ positive mDCs, including a DC2 subset (expressing *FCGR2B*, *CLEC10A*, and *CD1E*) and an inflammatory DC3 subset (expressing *CD14*, *CD163*, and *S100A8/9*)^14, 15, 16, 17^. Furthermore, multiple studies have described a separate mDC state known as “mature DCs enriched in immunoregulatory molecules” (mregDC cells), also referred to as LAMP3+ DCs, characterized by expression of *LAMP3*, *CCR7*, and *BIRC3*^14, 18, 19^. Other proposed mDC subsets, including monocyte-like DC4 cells and AXL+ SIGLEC6+ DC5 cells, are not as well characterized or agreed-upon^15, 16^. Finally, plasmacytoid dendritic (pDC) cells are a well-described DC subset that is thought to be of lymphoid origin^14^.

Prior reports have described the presence of mDCs in IBM patients^7, 20, 21, 22^. Greenberg et al. first reported the presence of CD1C+ mDCs (cDC2 cells) in the muscle of IBM patients by immunohistochemistry^20^. This was followed by another study showing CCR7+ CD1C+ mDCs (markers of cDC2 and mregDC cells) in IBM muscle by immunohistochemistry^21^. Most recently, single-nucleus RNA-sequencing (snRNA-seq) of IBM muscle revealed the presence of two clusters of mDCs, corresponding to cDC1 cells and mregDC cells, at higher proportions than in muscle of IMNM and non-myositis control patients^22^. We also recently showed that *XCR1*, an established cDC1 marker, and its T cell-derived ligands *XCL1* and *XCL2*, are all specifically differentially overexpressed in muscle of IBM patients compared to that of patients with other inflammatory myopathies^10^. Nevertheless, there has not been a systematic and comprehensive analysis of the main mDC subsets in IBM, nor has there been a study correlating these subsets with markers of disease activity in IBM. In this study, we analyzed snRNA-seq data from three datasets for the presence and characteristics of cDC1 cells, cDC2 cells, and mregDC cells in skeletal muscle of IBM and other myositis and control patients. We then analyzed bulk RNA-sequencing (RNA-seq) data from two datasets to correlate specific markers of these subsets with markers of muscle damage, immune cells, and type 1 inflammation in skeletal muscle of IBM and other myositis and control patients.

## RESULTS

### Initial clustering of single-nucleus RNA-sequencing data

The composition of the three datasets included in the snRNA-seq analysis is included in Table 1, with datasets B and C being publicly available^22, 23^. Before quality control, there were 318,698 unique cells included across the three datasets. After quality control, 233,026 cells (73%) were maintained for transformation. UMAP clustering revealed the unnamed clusters shown in Supplementary Figure 1A. We named eight clusters based on expression of specific markers, shown in Figure 1A and B and Supplementary Figure 2. These included myeloid cells, lymphocytes, endothelial cells, smooth muscle cells, skeletal muscle progenitors, myonuclei, fibroblasts, and adipocytes. The lymphocyte cluster was further subclustered into B cells, plasma cells, and T & NK cells (Supplementary Figures 3-4). Notably, the majority of B cells (69%), plasma cells (75%), T & NK cells (71%), and myeloid cells (65%) were from IBM samples (Figure 1C).

**Figure 1.**
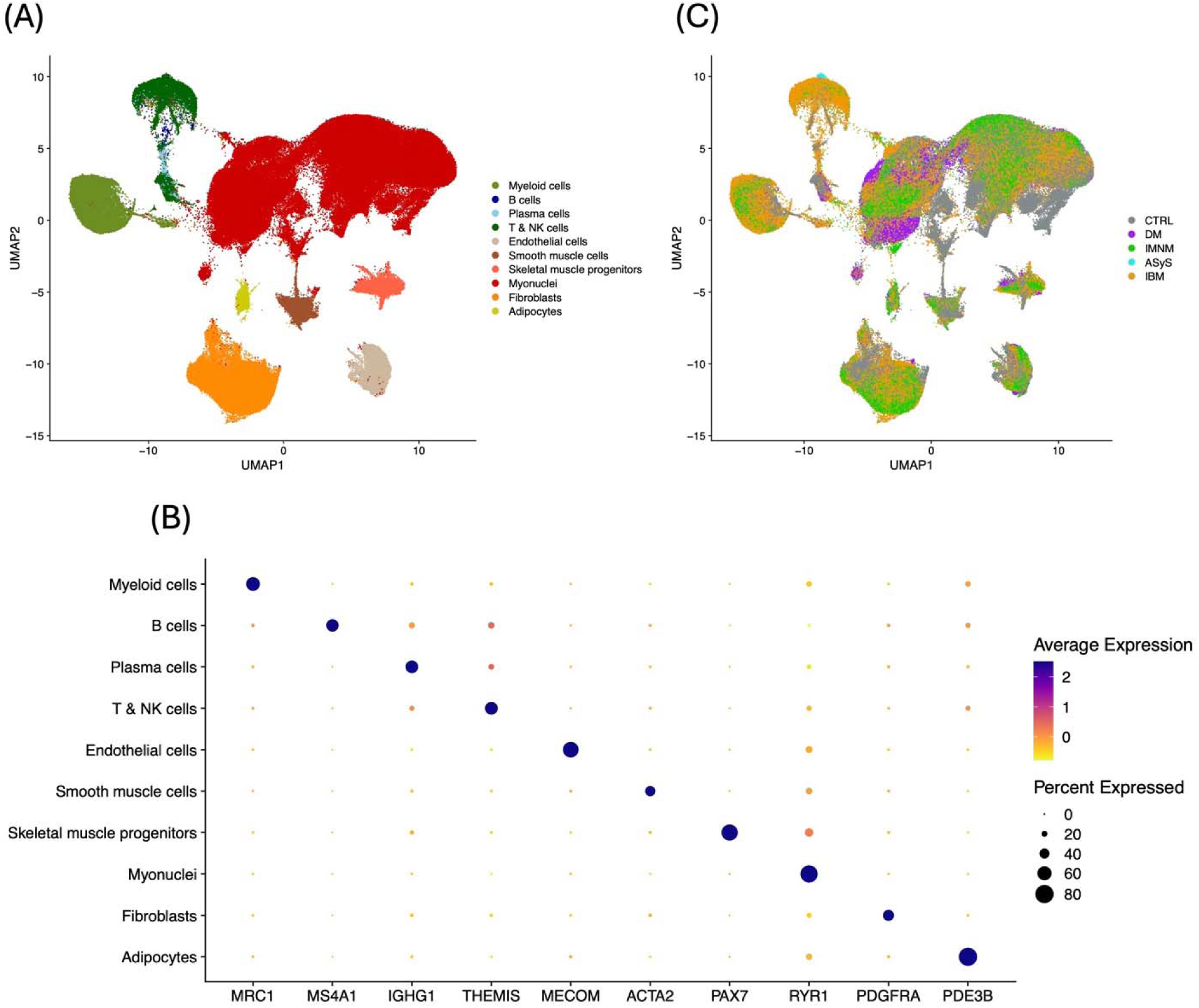
Clustering of all filtered cells and representative genes from snRNA-seq. (A) Label assignments of cell clusters. (B) Representative markers expressed in each cell cluster. (C) Clinical groups of cells.

**Table 1.**
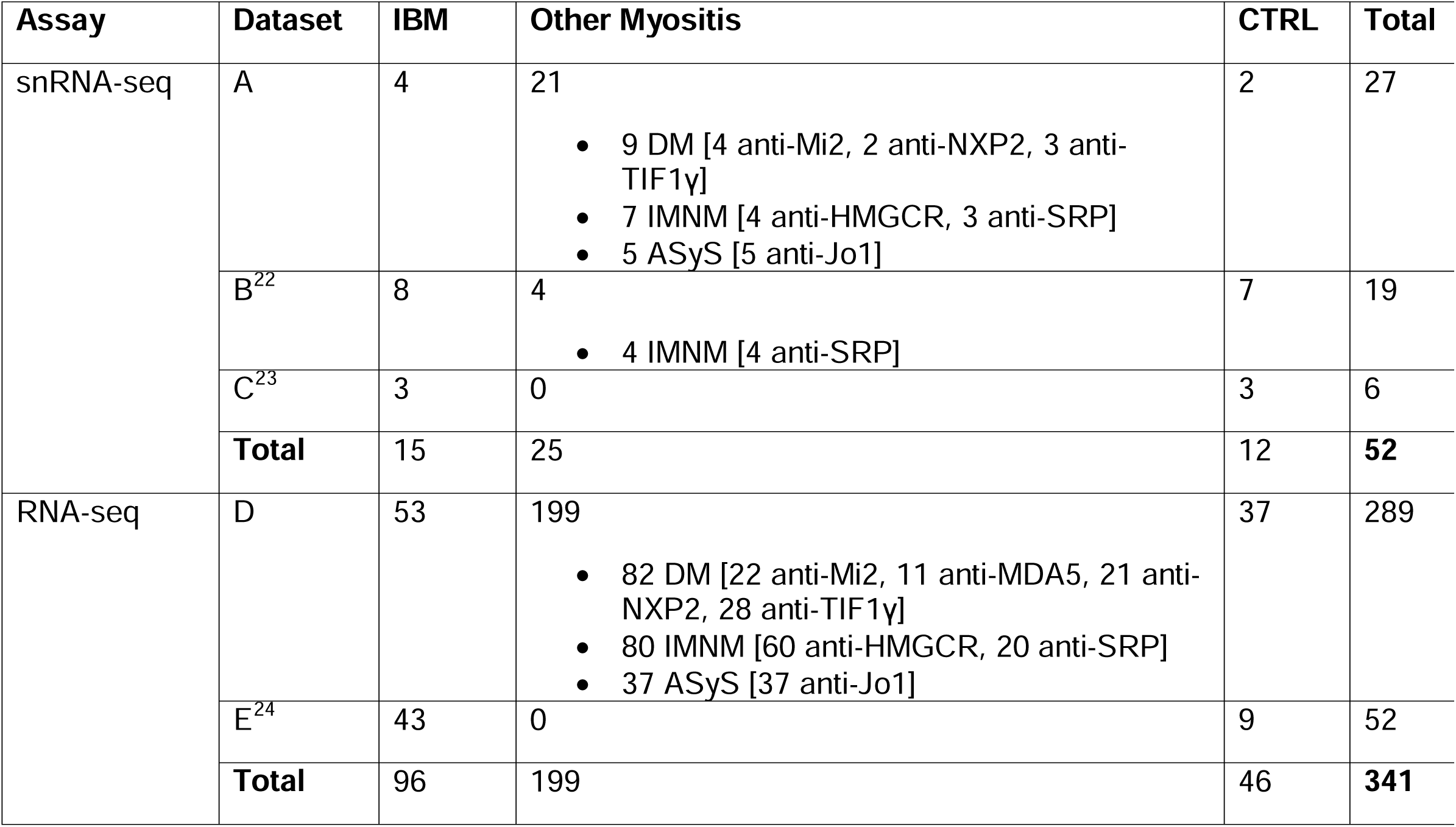
Patient composition of datasets used in snRNA-seq and RNA-seq analyses. Datasets A and D represented patients from our own cohort, who underwent snRNA-seq and RNA-seq of skeletal muscle, respectively. Datasets B and C were publicly availably snRNA-seq datasets of skeletal muscle, while Dataset E was a publicly available RNA-seq dataset of skeletal muscle^22, 23, 24^.

### Identification and quantification of myeloid dendritic cells in single-nucleus RNA-sequencing data

Clustering of the 17,245 myeloid cells revealed three main distinguishable clusters: two corresponding to mDC subsets and a larger “Other Myeloid cells” cluster, mainly made up of monocytes and macrophages (Figure 2A, Supplementary Figure 5A). Specifically, 575 cells made up a cluster that corresponded to cDC1 markers and 189 cells made up a cluster that corresponded to mregDC markers, with the rest in the “Other Myeloid cells” cluster. Since we were not able to identify a cDC2 cluster by unsupervised clustering, we labeled cDC2 cells manually as *CD1C*+ cells in the “Other Myeloid cells” cluster lacking expression of the specific monocyte marker *C5AR1*^17^. This revealed 277 cDC2 cells that largely clustered in one region of the UMAP, near the mregDCs (Figure 2A).

**Figure 2.**
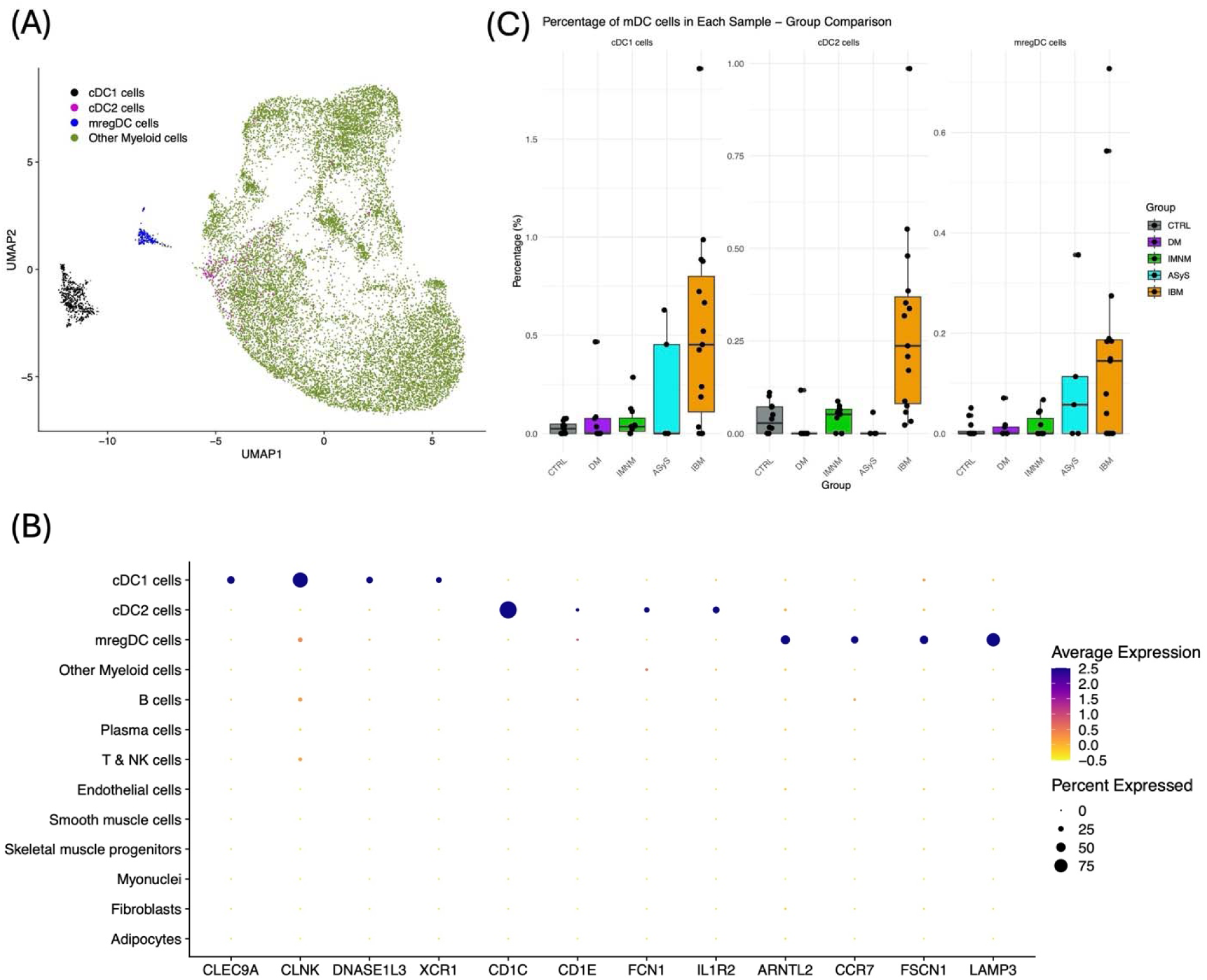
Clustering of Myeloid cells and proportions of mDCs by group from snRNA-seq. (A) Label assignments of myeloid cells. (B) Representative genes expressed in each mDC subtype. (C) Proportions of mDCs in each sample among all cells.

Of the 359 genes identified as specific to the cDC1 cell cluster, at least 49 were previously reported cDC1 markers (Supplementary File, Extended Data Figure 1A)^13, 15^. Of these, we selected 4 markers (*CLEC9A*, *CLNK*, *DNASE1L3*, *XCR1*) with strong specificity for the cDC1 cell cluster for our RNA-seq analysis (Figure 2B, Extended Data Figure 2).

Of the 236 genes identified as specific to the cDC2 cell cluster, at least 22 were previously reported cDC2 markers (Supplementary File, Extended Data Figure 1B). These included previously published markers of DC2 cells (e.g., *CD1E, FCGR2B*), DC3 cells (e.g., *CIITA*, *ITGAM*), and both (e.g., *CD1C*, *FCN1*)^15, 16^. Given these mixed markers, and the lack of clear subclustering of cDC2 cells into these two subsets, we maintained cDC2 cells as one group. Among the cDC2-specific genes was also the decoy interleukin receptor *IL1R2*, which, although not previously published as a cDC2 marker, was remarkably specific for the cluster compared to most published markers and expressed in both IBM and non-IBM samples. As such, we selected *CD1C*, *CD1E*, *FCN1*, and *IL1R2* as especially specific for the cDC2 cell cluster for our RNA-seq analysis (Figure 2B, Extended Data Figure 2).

Of the 423 genes identified as specific to the mregDC cell cluster, at least 96 were previously reported mregDC markers (Supplementary File, Extended Data Figure 1C)^14, 18, 19^. Of these, we selected 4 markers (*ARNTL2*, *CCR7*, *FSCN1*, *LAMP3*) with strong specificity for the mregDC cell cluster for our RNA-seq analysis (Figure 2B, Extended Data Figure 2).

All three mDC subsets were proportionally increased among all cells of IBM samples compared to all cells of other myositis and control (CTRL) samples (Figure 2C), with cDC1 cells being the most abundant of the three. There was notable variation of mDC levels among IBM samples, with cDC1 levels correlating positively with mregDC levels (Extended Data Figure 3).

### Evidence for mDC involvement in IFN-II response and CD8+ T cell activation in single-nucleus RNA-sequencing data

We next examined differential expression of 58 inflammatory genes in mDC cells derived from IBM patients compared to those derived from non-IBM patients. These included the genes encoding the IFN-γ receptor (*IFNGR1* and *IFNGR2*), five IFN-II-inducible genes, the genes encoding ligands of KLRG1 (*CDH1* and *CDH2*), 11 co-stimulatory genes, nine co-inhibitory genes, 19 human leukocyte antigen (HLA) genes encoding major histocompatibility complex (MHC) proteins, and ten genes encoding cytokines typically secreted by myeloid cells (Extended Data Table 1).

Of the seven IFN-II-related genes, both *IFNGR1* and *GBP2* were among the 236 cDC2-specific genes we previously identified, while *IFI30* was also most expressed in cDC2 cells. *IFNGR2* was among the 423 mregDC-specific genes, with cDC2 cells also exhibiting high expression of this gene (Supplementary File, Extended Data Figure 4). These observations support the notion that cDC2 cells are principal responders to T cell-derived IFN-γ. Furthermore, our differential expression analysis revealed that cDC2 cells from IBM patients generally had increased expression of IFN-II-related genes compared to cDC2 cells from non-IBM patients, although this did not reach statistical significance (Extended Data Table 1).

Interestingly, 6 MHC-II-encoding genes (*HLA-DMA*, *HLA-DMB*, *HLA-DQA1*, *HLA-DQB1*, *HLA-DRB1*, *HLA-DRB5*) and the co-stimulatory gene *CD86* were also among the 236 cDC2-specific genes, validating this cell type’s role as an activator of CD4+ T cells (Supplementary File, Extended Data Figure 4). In IBM patients, cDC2 cells appeared to express higher levels of multiple co-stimulatory and HLA genes than in non-IBM patients, but this did not meet statistical significance (Extended Data Table 1).

On the other hand, the co-stimulatory gene *CD80*, the immune checkpoints *CD274* (PD-L1) and *PDCD1LG2* (PD-L2), and *IL15* were among the 423 mregDC-specific genes, with the latter three being established mregDC markers (Supplementary File, Extended Data Figure 4)^14, 18, 19^. In IBM patients, mregDC cells expressed significantly higher levels of *IL15* than in non-IBM patients and trended towards increased expression of other co-stimulatory, co-inhibitory, and HLA genes without reaching statistical significance (Extended Data Table 1).

Finally, cDC1 cells from IBM patients expressed higher levels of the co-stimulatory gene *CD40*, the co-inhibitory gene *HAVCR2*, seven HLA genes, as well as *IL15* than those from non-IBM patients, representing the most substantial activation among mDC subsets (Extended Data Table 1). Interestingly, we observed that cDC1 cells expressed higher average amounts of both KLRG1 ligands *CDH1* (E-cadherin) and *CDH2* (N-cadherin) than other cell types, with *CDH2* being among the 359 cDC1-specific genes we previously identified (Figure 3A, Supplementary File, Extended Data Figure 4). Furthermore, cDC1 cells from IBM samples appeared to have increased expression of both *CDH1* and *CDH2* compared to cDC1 cells from non-IBM samples, and to a larger degree than for other mDC subsets, although this did not reach statistical significance after adjustment for multiple comparisons (Extended Data Table 1, Extended Data Figure 5). In addition, *CLEC9A* expression in myeloid cells of IBM samples was significantly correlated with *KLRG1* expression in T & NK cells of those samples, and to a greater extent (by ρ) than *CD1C* and *LAMP3* (Figure 3B). This correlation was notably absent in non-IBM samples (Extended Data Figure 6A). Taken together, these findings implicate cDC1 cells as principal activators of KLRG1+ CD8+ T cells in IBM.

**Figure 3.**
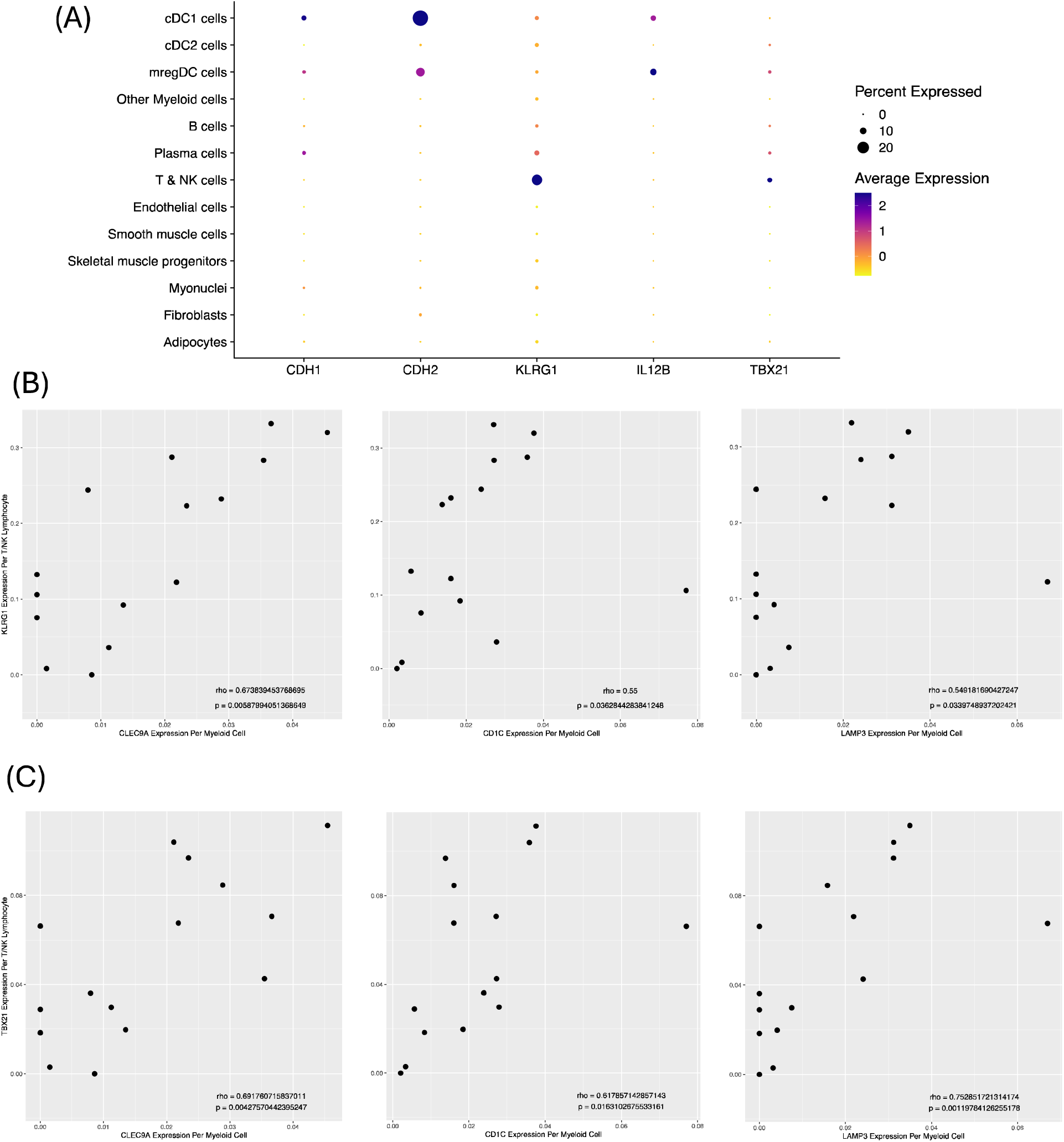
Correlation of mDC markers with Inflammatory T cell markers in IBM samples from snRNA-seq. (A) Expression of *KLRG1* and its ligands (*CDH1*, *CDH2*), as well as *TBX21* and its stimulating cytokine (*IL12B*) in each cell type among IBM samples. (B) Correlation of *KLRG1* expression per T/NK cell with *CLEC9A*, *CD1C*, and *LAMP3* expression per myeloid cell in IBM samples. (C) Correlation of *TBX21* expression per T/NK cell with *CLEC9A*, *CD1C*, and *LAMP3* expression per myeloid cell in IBM samples.

Also notable was the expression of *IL12B*, encoding a portion of the TBX21+ T cell-activating cytokine IL-12, which appeared to be restricted to cDC1 and mregDC cells (Figure 3A, Extended Data Figure 5). The expression appeared markedly increased in cDC1 cells from IBM patients compared to cDC1 cells from non-IBM patients by log-fold change, although this was not statistically significant after adjustment for multiple comparisons (Extended Data Table 1). *CLEC9A*, *CD1C*, and *LAMP3* expression in myeloid cells of IBM samples were all significantly correlated with *TBX21* expression in T & NK cells of those samples, although the correlation was strongest for *LAMP3* and *CLEC9A* (Figure 3C). This correlation was notably absent in non-IBM samples (Extended Data Figure 6B). Altogether, these results implicate cDC1 cells and mregDC cells as activators of TBX21+ CD8+ T cells in IBM.

### Expression of mDC markers and correlation with markers of IBM disease activity in bulk RNA-sequencing data

The composition of the two datasets included in the RNA-seq analysis is included in Table 1. In our RNA-seq dataset (Dataset D), IBM samples had higher median expression of all four cDC1-specific genes than DM, IMNM, ASyS, and CTRL samples, in line with our snRNA-seq data (Figure 4A). IBM samples also had higher median expression of some cDC2- and mregDC-specific genes (*CD1C*, *CD1E*, and *CCR7*), while most other genes examined had similar expression between groups (Figure 4A). In the publicly available dataset (Dataset E)^24^, IBM samples had higher median expression of all mDC-specific genes compared to CTRL samples, but this was most prominent for mregDC-specific genes, followed by cDC1-specific genes (Figure 4B). Correlation analysis of specific mDC genes with each other revealed that, in IBM patients, all three mDC subsets were correlated with each other (Extended Data Figure 7).

**Figure 4.**
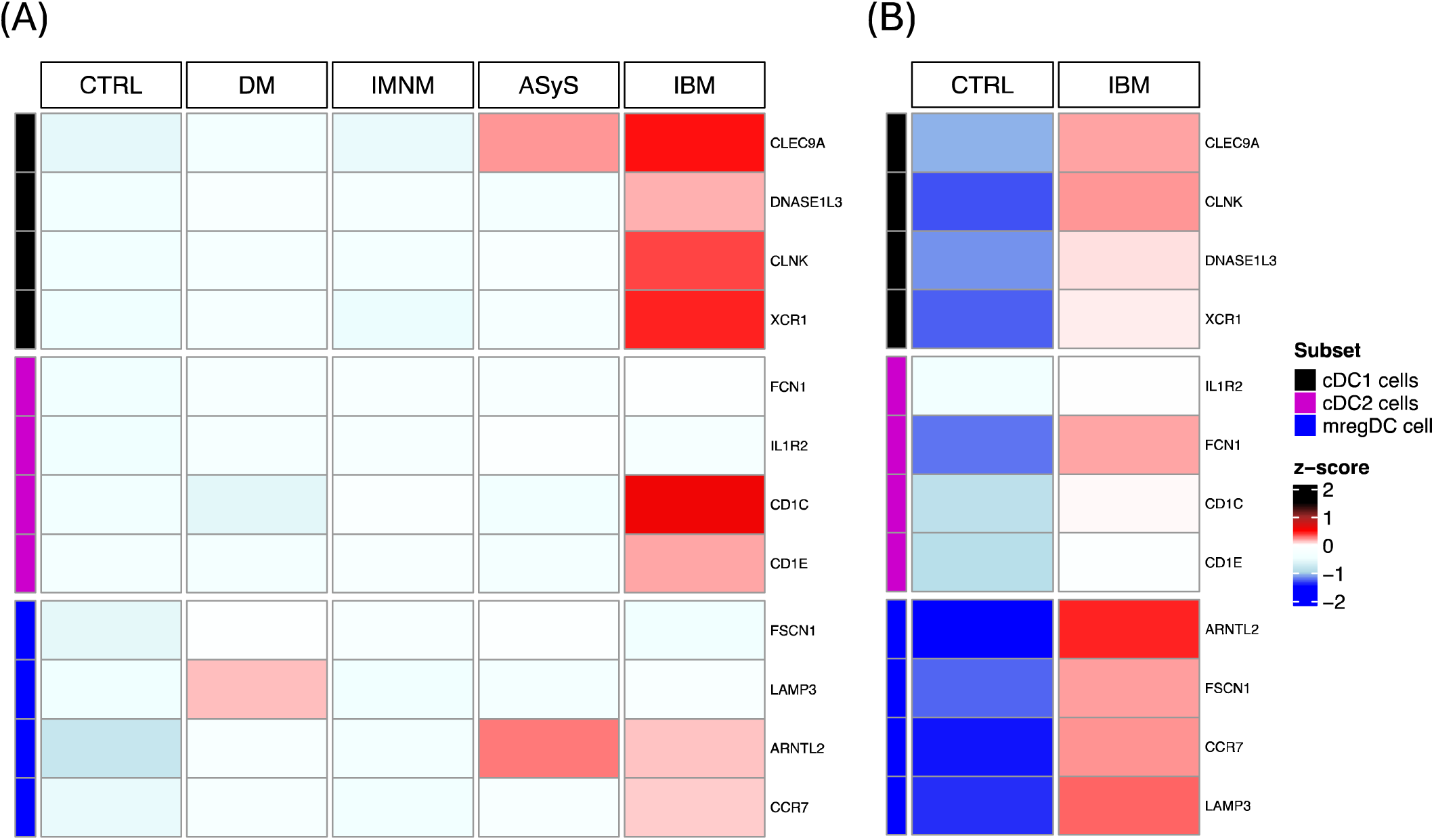
Median Expression Heatmaps of specific mDC genes in each group from RNA-seq. (A) Median Expression in each group in RNA-seq dataset D. (B) Median Expression in each group in RNA-seq dataset E.

We next performed correlation analysis of specific mDC genes with transcriptomic markers of muscle damage and regeneration, the IFN-II pathway, and immune cells (Figure 5). In Dataset D, mDC-specific genes from all three subsets were generally more strongly correlated with markers of the IFN-II pathway, T cell markers (including *TBX21* and *KLRG1*), and macrophages for IBM patients than other myositis and CTRL patients (Figure 5A-C). In Dataset E, cDC1-specific genes were more strongly correlated with these inflammatory markers for IBM patients than CTRL patients, while for the cDC2-specific and mregDC-specific genes the difference was less clear (Figure 5D-E).

**Figure 5.**
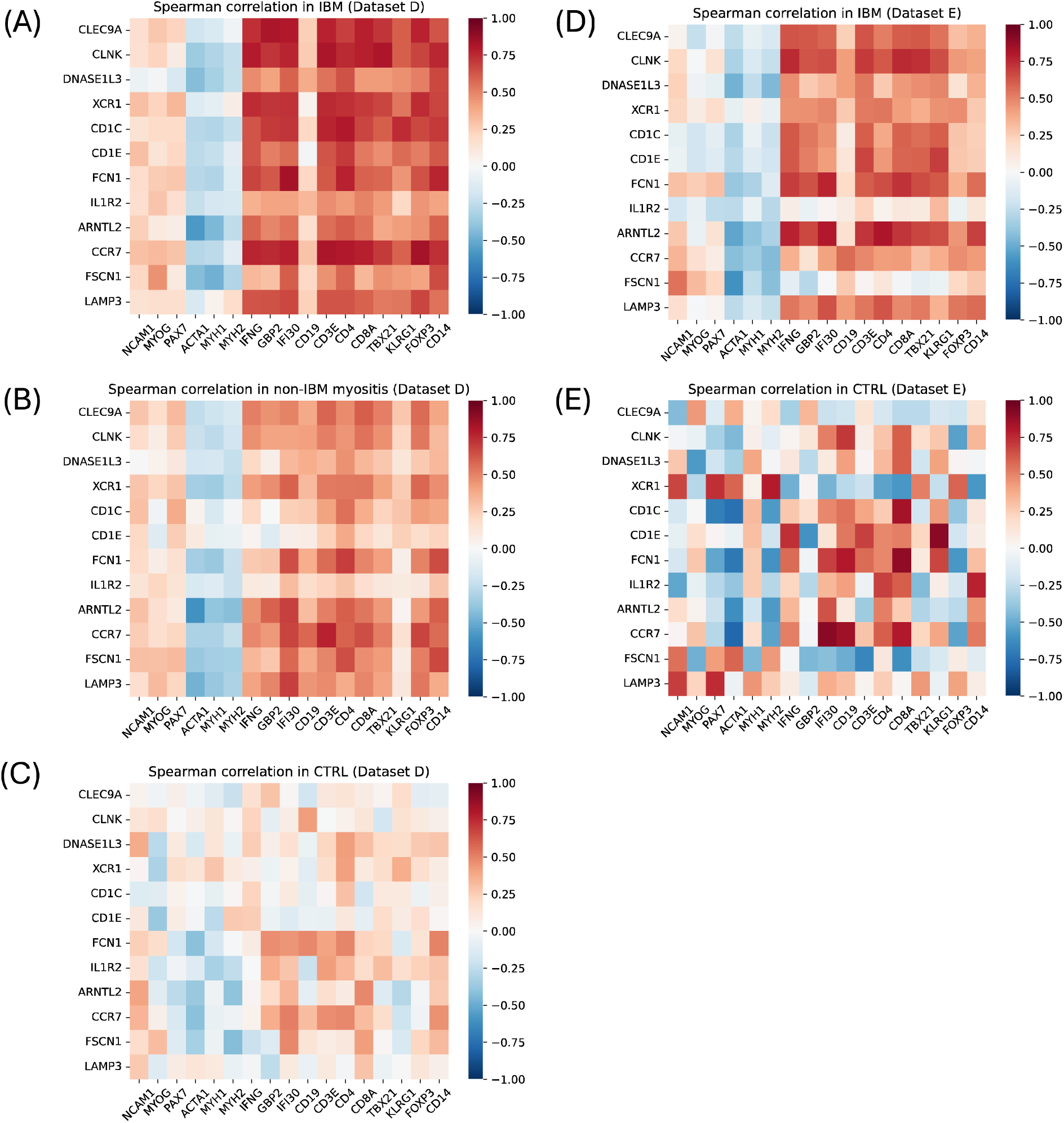
Correlation Heatmaps of specific mDC markers with disease markers in IBM, other myositis, and CTRL samples from RNA-seq. Correlation of expression of cDC1-specific genes (*CLEC9A*, *CLNK*, *DNASE1L3*, *XCR1*), cDC2-specific genes (*CD1C*, *CD1E*, *FCN1, IL1R2*), and mregDC-specific genes (*ARNTL2*, *CCR7, FSCN1*, *LAMP3*) versus markers of muscle regeneration (*NCAM1*, *MYOG*, *PAX7*), mature muscle markers (*ACTA1*, *MYH1*, *MYH2*), IFN-II pathway genes (*IFNG*, *GBP2*, *IFI30*), B cell marker (*CD19*), T cell markers (*CD3E*, *CD4*, *CD8A*, *TBX21*, *KLRG1*, *FOXP3*), and macrophage marker (*CD14*). Correlations shown for IBM samples in dataset D (A), non-IBM myositis samples in dataset D (B), CTRL samples in dataset D (C), IBM samples in dataset E (D), and CTRL samples in dataset E (E).

In order to discover additional IBM-specific genes correlated with each mDC subset in our RNA-seq data, we found differentially correlated genes in IBM with *CLEC9A*, *CD1C*, and *LAMP3* (Extended Data Figure 8). As expected, *CLEC9A* was differentially correlated with multiple cytotoxic markers (e.g., *CD8A*, *CD8B*, *FCGR3A*, *GZMB*, *GZMH*, *KLRK1*, *TBX21*), as well as IFN-II pathway genes (e.g., *CXCL9*, *CXCL10*, *CXCR3*, *GBP1*, *GBP2*, *IFI30*, *IFNG*, *IL12B*, *PSMB8*), two MHC-I-encoding HLA genes (*HLA-B* and *HLA-F*), and eight MHC-II-encoding HLA genes in IBM samples. On the other hand, *CD1C* was differentially correlated with *CD4*, several IFN-II pathway genes (e.g., *CCR5*, *CXCL9*, *CXCR3*, *IFI30*), ten MHC-II-encoding HLA genes, as well as *KLRB1* and *KLRG1* in IBM samples. *LAMP3* had the least differentially correlated genes in IBM samples, but they included *IL12B*, *CD28*, and *CTLA4*.

### Presence and correlations of mDC subsets in PM disease groups in bulk RNA-sequencing data

Given the common theme of CD8+ T cell muscle invasion of PM and IBM^25^, we sought to examine mDC-specific gene expression and correlation in two PM groups obtained from publicly available RNA-seq datasets^24, 26^. These were labeled as PM with CD8+ T cell invasion of MHC-I+ muscle (PM-CD8)^24^ and PM with mitochondrial pathology (PM-Mito)^26^. Interestingly, both groups exhibited high median expression of cDC1-specific genes, at equal or higher levels than IBM patients. Both groups also exhibited high expression of cDC2-specific genes, while PM-Mito patients also exhibited high expression of mregDC-specific genes (Extended Data Figure 9A). In both, cDC1-specific genes appeared to have stronger correlations with inflammatory markers (including *TBX21*, and, in PM-CD8, *KLRG1*) than in IBM patients, and, in PM-Mito patients, mregDC-specific genes also appeared to have stronger correlations with inflammatory markers than in IBM patients (Extended Data Figure 9B-C). This suggests a common role for mDCs in IBM and PM pathophysiology.

## DISCUSSION

While prior reports have described individual mDC subsets in IBM muscle, our results expand on these by providing a comprehensive analysis of three different types of mDCs in IBM patients compared to the other main types of myositis^7, 20, 21, 22^. Specifically, we have shown that cDC1 cells, cDC2 cells, and mregDC cells are present at higher proportions in IBM samples compared to other myositis and CTRL samples.

Although some IBM patients had low numbers of mDCs, cDC1 and mregDC presence was correlated among IBM samples in our snRNA-seq analysis. In the bulk RNA-seq analysis, specific markers of all three subsets were correlated with each other. We also observed IBM-specific mDC activation, especially in cDC1 cells, in our snRNA-seq analysis, as signified by increased expression of co-stimulatory genes, HLA genes, and cytokines.

Of the three, cDC1 cells made up the predominant mDC subset. Interestingly, cDC1 cells were the principal expressors of *CDH1* and *CDH2*, the two ligands of *KLRG1*, a marker of highly differentiated CD8+ cytotoxic T cells implicated in IBM^7^. CDH1 has been reported to be increased in IBM muscle by RNA-seq and immunohistochemistry, but the cellular source was not established^24^. Expression of both of these genes appeared higher in cDC1 cells from IBM samples compared to cDC1 cells from non-IBM samples. Furthermore, in our snRNA-seq analysis, cDC1 marker expression in myeloid cells was positively correlated with *KLRG1* expression in T & NK cells from IBM samples, something that was not true when using all myositis and CTRL samples. Taken together, these data suggest that, in IBM, cDC1 cells are induced to expand and express *CDH1* and *CDH2*, subsequently activating KLRG1+ CD8+ T cells. Indeed, the function of cDC1 cells in activating IFN-γ-secreting KLRG1+ cytotoxic T cells has been previously established in pancreatic cancer^27^. Moreover, although *CDH1* expression has been reported in immunoregulatory subtypes of DCs (and we observed some expression in mregDCs), it has also been reported in a pro-inflammatory mDC subtype in a mouse model of chronic T cell-mediated colitis^28, 29, 30^.

The other subtype of CD8+ T cell previously implicated in IBM is TBX21+ T_C_1 cells, which secrete IFN-γ and mediate type 1 inflammation^11^. *TBX21* has also been found to be highly co-expressed with *KLRG1* in human tissue, suggesting the presence of TBX21+ KLRG1+ T_C_1 cells^7^. Here, we showed that cDC1 marker expression in myeloid cells was positively correlated with *TBX21* expression in T & NK cells from IBM samples, but not in non-IBM samples. Additionally, the cDC1 marker *CLEC9A* was differentially correlated with *TBX21* in IBM samples by RNA-seq, in contrast to *CD1C* and *LAMP3*. We also showed that cDC1 cells from IBM patients, as well as mregDC cells, were principal expressors of *IL12B*, encoding part of the T_C_1-stimulating cytokine IL-12, and appeared to express it at higher levels than cDC1 cells from non-IBM patients. This provides further evidence of the role of cDC1 cells in initiating CD8+ T cell-mediated type 1 inflammation in IBM. This role of cDC1 cells in type 1 inflammation has previously been demonstrated in other inflammatory contexts, including white adipose tissue in obesity and renal tissue in late-stage anti-glomerular basement membrane disease ^31, 32^.

Interestingly, in two groups of PM patients, we also saw very high expression of cDC1-specific genes in RNA-seq data, with even stronger correlations with inflammatory genes, including *TBX21* and *KLRG1*, than in IBM patients. This suggests that cDC1 cells may be even more active in these patients and contribute to CD8+ T cell-mediated inflammation, underlying a similar pathophysiology as IBM. This, along with the fact that other mDC subset-specific genes were also overexpressed in these groups, adds to previous suggestions that some PM patients (including the PM-Mito group we examined) may represent early or atypical forms of IBM^26, 33, 34^.

Our snRNA-seq data also implicates cDC2 cells as principal responders to IFN-γ in IBM, via upregulation of IFN-II-inducible genes, and as activators of CD4+ T cells via specific expression of multiple HLA genes encoding MHC-II, and differential correlation of *CD1C* with *CD4* via RNA-seq. As a result, we hypothesize that they contribute to a positive feedback loop of type 1 inflammation, involving KLRG1+ TBX21+ T_C_1 cells and T_H_1 cells. On the other hand, the role of mregDC cells in IBM is less clear, but they appear to exhibit immunoregulatory features consistent with their published function, including specific expression of the immune checkpoint genes *CD274* (PD-L1) and *PDCD1LG2* (PD-L2). Given the evidence that cDC1 and cDC2 cells can acquire the mregDC “state”, further study is warranted to assess for such a transition in IBM and the role of the cDC1-mregDC balance in phenotype and response to immunosuppression^14^.

Our study has several limitations. Although we included samples from other myositis samples for our comparisons, we did not have PM samples in our snRNA-seq datasets and some myositis groups (e.g., overlap myositis) were not included in either of our analyses. To analyze our RNA-seq data, we used highly specific mDC markers identified from snRNA-seq analysis, but this nevertheless assumes that the same cell types are present and exhibit the same marker expression in the RNA-seq samples. Finally, although we provide suggestive evidence of the roles of cDC1, cDC2, and mregDC cells in modulating inflammation in IBM, this requires additional functional assays to confirm.

In conclusion, we identified the increased presence and activation of three mDC subsets, cDC1 cells, cDC2 cells, and mregDC cells, in skeletal muscle of IBM patients and their positive correlation with markers of the IFN-II pathway and T cell subsets.

Furthermore, we propose cDC1 cells to be principal activators of KLRG1+ and TBX21+ CD8+ T cells in IBM via upregulation of *IL12B* and the KLRG1 ligands *CDH1* and *CDH2*. We also propose cDC2 cells to be principal responders to IFN-γ via upregulation of IFN-II-inducible genes and principal activators of CD4+ T cells via specific expression of multiple MHC-II-encoding genes. Finally, we also identified increased mDC-specific gene expression and correlation with inflammatory genes in two groups of PM patients, suggesting a common pathophysiological theme with IBM patients.

## ONLINE METHODS

### Patients

The patients whose skeletal muscle samples were used in our analyses are summarized in Table 1. For the snRNA-seq analysis, our dataset (Dataset A) included 4 IBM patients, 9 DM patients, 7 IMNM patients, 5 ASyS patients, and 2 control (CTRL) patients. We also analyzed two publicly available snRNA-seq datasets: Dataset B, including 8 IBM patients, 4 IMNM patients, and 7 CTRL patients^22^, and Dataset C, including 3 IBM patients and 3 CTRL patients^23^. For the RNA-seq analysis, our dataset (Dataset D) included 53 IBM patients, 82 DM patients, 80 IMNM patients, 37 ASyS patients, and 37 CTRL patients. We also analyzed one publicly available RNA-seq dataset, Dataset E, including 43 IBM patients and 9 CTRL patients^24^. Additionally, we performed a supplementary RNA-seq analysis where we also included two groups of PM patients from two publicly available datasets: 6 patients labeled as PM with CD8+ T cells invading non-necrotic MHC-I+ muscle (PM-CD8)^24^, and 7 patients with PM with mitochondrial pathology (PM-Mito)^26^.

Muscle biopsies for our datasets (Datasets A and D) were obtained from institutional review board-approved longitudinal cohorts from the National Institutes of Health in Bethesda, MD; the Johns Hopkins Myositis Center in Baltimore, MD; the Vall d’Hebron Hospital and the Clinic Hospital in Barcelona, Spain; and the Charité-Universitätsmedizin in Berlin, Germany. Patients were classified as IBM if they fulfilled Lloyd’s criteria for inclusion body myositis^35^. Patients were classified as DM, IMNM, or ASyS if they fulfilled the Casal and Pinal criteria and tested positive for the following myositis specific autoantibodies: anti-Mi2 (DM), anti-MDA5 (DM), anti-NXP2 (DM), anti-TIF1γ (DM), anti-HMGCR (IMNM), anti-SRP (IMNM), or anti-Jo1 (ASyS)^36^. Autoantibody testing was performed using one or more of the following techniques: ELISA, immunoprecipitation of proteins generated by *in vitro* transcription and translation (IVTT-IP), line blotting (EUROLINE myositis profile), or immunoprecipitation from 35S-methionine-labeled HeLa cell lysates.

### Standard protocol approvals and patient consent

This study was approved by the Institutional Review Boards of the National Institutes of Health, the Johns Hopkins University, the Clinic Hospital, the Vall d’Hebron University Hospital, and the Charité-Universitätsmedizin Berlin. Written informed consent was obtained from each participant. All methods were performed in accordance with the relevant guidelines and regulations.

### Single-nucleus RNA-sequencing

snRNA-seq was performed on frozen muscle biopsy specimens as previously described^37, 38^. Briefly, frozen muscle samples were homogenized and underwent a sucrose-gradient ultracentrifugation nuclei isolation protocol adapted from Schirmer et al^39^. The nuclei were counted using a manual hemocytometer and between 2000 and 3000 nuclei per sample were loaded into the 10X Genomic Single-Cell 3′ system.

### Bulk RNA-sequencing

RNA-seq was performed on frozen muscle biopsy specimens as previously described^8, 40, 41, 42, 43, 44, 45^. Briefly, muscle biopsies underwent immediate flash freezing and were stored at -80°C across all contributing centers. Samples were then transported in dry ice to the NIH and processed uniformly to prepare the library and conduct the analysis. RNA was extracted with TRIzol. Libraries were either prepared with the NeoPrep system according to the TruSeq Stranded mRNA Library Prep protocol (Illumina, San Diego, CA) or with the NEBNext Poly(A) mRNA Magnetic Isolation Module and Ultra^™^ II Directional RNA Library Prep Kit for Illumina (New England BioLabs, ref. #E7490, and #E7760).

### Statistical and bioinformatic analysis

snRNA-seq reads from Dataset A were demultiplexed and aligned using cellranger/6.0.1. Seurat/5.1.0 and R/4.4.1 was used for snRNA-seq analysis. Specifically, Datasets A, B, and C were merged into one Seurat object using the merge and JoinLayers functions. Quality control was performed to exclude cells with ≤200 or ≥3000 features, ≥10% mitochondrial or ribosomal features, or ≥1% hemoglobin or platelet features. This was followed by normalization using SCTransform (regressing out the dataset and sample ID), integration using RunHarmony, UMAP generation, and cluster identification using FindClusters. Cell clusters were named based on gene expression patterns. This process was repeated to further subcluster the lymphocyte and myeloid cell clusters. As a note, cDC2 cells were not identified by unsupervised clustering, and were instead manually selected from the “Other Myeloid cells” cluster based on having >0 SCT expression of the cDC2 marker *CD1C* and 0 SCT expression of the specific monocyte marker C5AR1^17^. Markers of each mDC subset were identified using the FindMarkers function, by selecting those genes overexpressed in the subset compared to all cells (adjusted p-value <0.001), all myeloid cells (adjusted p-value <0.001) and each other individual cell type (p-value <0.05). In addition, differential expression of 58 inflammatory genes in each mDC subset in IBM versus non-IBM patients was assessed using the FindMarkers function (adjusted p-value <0.05). Spearman’s rank correlations were performed to correlate mDC marker expression per myeloid cell to each other and to *KLRG1* and *TBX21* expression per T & NK cell across samples (p-value <0.05). All snRNA-seq visualizations were created in R using functions contained in Seurat, scCustomize/3.0.1, and ggplot2/3.5.1. Adjusted p-values were determined by Bonferroni correction for multiple gene comparisons.

For our RNA-seq analysis (Dataset D), sequencing reads were demultiplexed using bcl2fastq/2.20.0 and preprocessed using fastp/0.23.4. The abundance of each gene was determined using Salmon/1.5.2. For both Dataset D and Dataset E (analyzed separately), counts were normalized using the Trimmed Means of M values (TMM) from edgeR/4.2.1 for graphical analysis. Heatmaps, generated in R, were used to visualize median expression of mDC markers identified from snRNA-seq analysis in each myositis or CTRL group. Correlation heatmaps, generated in Python, were used to visualize Spearman’s rank correlations of these markers with each other, established markers of mature and regenerating muscle, the IFN-II pathway, B cells, T cell subsets, and macrophages. Spearman’s rank correlations were also used to correlate mDC markers with other genes in IBM and non-IBM samples from Dataset D. Differentially correlated genes in IBM patients were defined as genes correlated with these markers in IBM with ρ >0.7 (all adjusted p-values <0.001, by Benjamini-Hochberg correction), and with Δρ >0.2 compared to correlation in non-IBM samples.

## Acknowledgments

Julie Thompson for her invaluable help maintaining the NIH Natural History Protocol, the NIAMS Sequencing Core, and its members.

## Contributorship

All authors contributed to the development of the manuscript, including interpretation of results, substantive review of drafts, and approval of the final draft for submission.

## Ethical approval information

All biopsies were from subjects enrolled in institutional review board (IRB)-approved longitudinal cohorts in the National Institutes of Health, the Johns Hopkins, the Clinic Hospital, the Vall d’Hebron Hospital, and the Charité-Universitätsmedizin Berlin.

## Patient and public involvement

Patients and/or the public were not involved in the design, conduct, reporting, or dissemination plans of this research.

## Data availability statement

Any anonymized data not published within the article will be shared by request from any qualified investigator.

