## Extended Data Figures for "Activated Dendritic Cell Subsets Characterize Muscle of Inclusion Body Myositis Patients and Correlate with KLRG1+ and TBX21+ CD8+ T cells"

**Extended Data Table 1. Expression of inflammatory genes in mDC cells from IBM samples versus non-IBM samples.** Showing log2FC, adjusted p-value, and unadjusted p-value of differential expression of 58 inflammatory genes in cDC1 cells, cDC2 cells, and mregDC cells in IBM versus non-IBM samples. The 58 genes include: 7 genes in IFN-II pathway (*IFNGR1*, *IFNGR2*, *IFI30*, *GBP1*, *GBP2*, *PSMB8*, *CXCL9*), 11 co-stimulatory genes (*CD40*, *CD58*, *CD70*, *CD80*, *CD86*, *ICOSLG*, *TNFSF4*, *TNFSF9*, *TNFSF14*, *TNFSF18*, *ICAM1*), 2 genes encoding ligands of KLRG1 (*CDH1*, *CDH2*), 9 co-inhibitory genes (*HAVCR2*, *TIMD4*, *CD274*, *PDCD1LG2*, *TNFRSF14*, *LILRB2*, *LILRB4*, *CD276*, *VTCN1*), 19 HLA genes, and 10 genes encoding cytokines (*IL1A*, *IL1B*, *IL6*, *IL10*, *IL12A*, *IL12B*, *IL15*, *IL18*, *IL23A*, *TNF*). Bolded values are statistically significant (adjusted p-value <0.05); green-colored values are overexpressed by p-value <0.05, while red-colored values are underexpressed by p-value <0.05.

| | **cDC1** | | | **cDC2** | | | **mregDC** | | |
|---|---|---|---|---|---|---|---|---|---|
| **Gene** | **Log2FC** | **Adjusted p-value** | **p-value** | **Log2FC** | **Adjusted p-value** | **p-value** | **Log2FC** | **Adjusted p-value** | **p-value** |
| IFNGR1 | 0.962 | 1.00E+00 | 1.04E-01 | NA | NA | NA | 1.313 | 1.00E+00 | 9.25E-02 |
| IFNGR2 | **2.196** | **8.50E-05** | **3.23E-09** | 1.432 | 9.47E-01 | 3.60E-05 | 1.550 | 1.00E+00 | 5.16E-04 |
| IFI30 | -0.139 | 1.00E+00 | 5.56E-01 | 0.776 | 1.00E+00 | 3.39E-01 | 0.605 | 1.00E+00 | 6.95E-01 |
| GBP1 | **-1.218** | **3.41E-02** | **1.29E-06** | 2.152 | 1.00E+00 | 2.42E-03 | 0.283 | 1.00E+00 | 9.31E-01 |
| GBP2 | -0.384 | 1.00E+00 | 2.21E-02 | 2.351 | 1.00E+00 | 6.04E-05 | 0.820 | 1.00E+00 | 7.05E-01 |
| PSMB8 | 1.879 | 1.00E+00 | 1.08E-02 | 0.269 | 1.00E+00 | 9.02E-01 | 0.235 | 1.00E+00 | 8.83E-01 |
| CXCL9 | 0.917 | 1.00E+00 | 3.20E-01 | 3.501 | 1.00E+00 | 9.47E-03 | 0.302 | 1.00E+00 | 5.00E-01 |
| CD40 | **2.908** | **1.07E-03** | **4.07E-08** | 2.031 | 1.00E+00 | 4.75E-02 | 2.008 | 1.00E+00 | 2.07E-02 |
| CD58 | 1.566 | 1.00E+00 | 1.21E-04 | 0.274 | 1.00E+00 | 3.58E-01 | 0.916 | 1.00E+00 | 1.10E-01 |
| CD70 | NA | NA | NA | NA | NA | NA | -0.542 | 1.00E+00 | 9.95E-01 |
| CD80 | 2.786 | 1.00E+00 | 3.32E-03 | 1.138 | 1.00E+00 | 2.59E-01 | 1.338 | 1.00E+00 | 1.32E-02 |
| CD86 | 0.700 | 1.00E+00 | 8.34E-02 | NA | NA | NA | 1.110 | 1.00E+00 | 2.08E-01 |
| ICOSLG | -1.191 | 1.00E+00 | 3.40E-01 | NA | NA | NA | -1.864 | 1.00E+00 | 5.69E-02 |
| TNFSF4 | -1.969 | 1.00E+00 | 1.40E-01 | NA | NA | NA | NA | NA | NA |
| TNFSF9 | -0.384 | 1.00E+00 | 2.79E-01 | NA | NA | NA | NA | NA | NA |
| TNFSF14 | -1.554 | 1.00E+00 | 1.41E-01 | NA | NA | NA | NA | NA | NA |
| TNFSF18 | NA | NA | NA | NA | NA | NA | NA | NA | NA |
| ICAM1 | 0.784 | 1.00E+00 | 4.39E-02 | 2.819 | 1.00E+00 | 5.34E-04 | 2.223 | 1.00E+00 | 1.48E-03 |
| CDH1 | 2.839 | 1.00E+00 | 1.06E-02 | NA | NA | NA | 1.235 | 1.00E+00 | 2.74E-01 |
| CDH2 | 2.001 | 5.21E-01 | 1.98E-05 | 0.331 | 1.00E+00 | 3.04E-01 | 1.008 | 1.00E+00 | 2.44E-01 |
| HAVCR2 | **1.591** | **2.25E-02** | **8.55E-07** | 0.988 | 1.00E+00 | 2.91E-02 | 0.913 | 1.00E+00 | 1.32E-01 |
| TIMD4 | 1.031 | 1.00E+00 | 2.14E-01 | 0.746 | 1.00E+00 | 3.04E-01 | -0.350 | 1.00E+00 | 5.37E-01 |
| CD274 | 3.523 | 1.00E+00 | 5.29E-03 | 1.746 | 1.00E+00 | 1.10E-01 | 2.142 | 1.00E+00 | 8.21E-04 |
| PDCD1LG2 | 0.491 | 1.00E+00 | 6.86E-01 | 1.424 | 1.00E+00 | 3.75E-02 | 2.136 | 1.00E+00 | 2.35E-03 |
| TNFRSF14 | 0.300 | 1.00E+00 | 7.41E-01 | 0.816 | 1.00E+00 | 1.18E-01 | 2.405 | 1.00E+00 | 8.68E-02 |
| LILRB2 | -0.232 | 1.00E+00 | 8.94E-01 | NA | NA | NA | 0.650 | 1.00E+00 | 1.00E+00 |
| LILRB4 | -0.232 | 1.00E+00 | 7.50E-01 | 1.889 | 1.00E+00 | 1.32E-02 | -0.542 | 1.00E+00 | 6.60E-01 |
| CD276 | -0.161 | 1.00E+00 | 4.43E-01 | -0.669 | 1.00E+00 | 3.60E-01 | -0.542 | 1.00E+00 | 8.41E-01 |
| VTCN1 | NA | NA | NA | NA | NA | NA | NA | NA | NA |
| HLA-A | 0.549 | 1.00E+00 | 3.15E-01 | 1.889 | 1.00E+00 | 2.56E-02 | 0.690 | 1.00E+00 | 1.56E-01 |
| HLA-B | -0.177 | 1.00E+00 | 5.40E-01 | 1.013 | 1.00E+00 | 1.13E-01 | NA | NA | NA |

| | | | | | | | | | |
|---|---|---|---|---|---|---|---|---|---|
| HLA-C | NA | NA | NA | 1.039 | 1.00E+00 | 8.61E-02 | -0.157 | 1.00E+00 | 9.97E-01 |
| HLA-DMA | 1.783 | 1.92E-01 | 7.29E-06 | 0.623 | 1.00E+00 | 6.41E-02 | 2.628 | 1.00E+00 | 1.09E-02 |
| HLA-DMB | **1.700** | **3.10E-02** | **1.18E-06** | 0.908 | 1.00E+00 | 1.06E-02 | 3.323 | 1.00E+00 | 1.35E-02 |
| HLA-DOA | 1.732 | 1.00E+00 | 6.02E-02 | -0.361 | 1.00E+00 | 8.55E-01 | 1.898 | 1.00E+00 | 8.68E-02 |
| HLA-DOB | 1.178 | 1.00E+00 | 1.60E-01 | -1.032 | 1.00E+00 | 9.81E-01 | 1.738 | 1.00E+00 | 1.15E-01 |
| HLA-DPA1 | **1.634** | **6.04E-08** | **2.29E-12** | -0.222 | 1.00E+00 | 7.82E-01 | 1.701 | 1.00E+00 | 5.36E-03 |
| HLA-DPB1 | **0.791** | **7.97E-06** | **3.03E-10** | 0.651 | 1.00E+00 | 6.72E-03 | 1.634 | 1.00E+00 | 1.05E-03 |
| HLA-DQA1 | **2.013** | **4.70E-11** | **1.79E-15** | 0.679 | 1.00E+00 | 5.41E-04 | 2.022 | 2.10E-01 | 7.99E-06 |
| HLA-DQA2 | 2.457 | 1.00E+00 | 7.95E-03 | -0.991 | 1.00E+00 | 8.75E-01 | 0.650 | 1.00E+00 | 4.29E-01 |
| HLA-DQB1 | **1.512** | **1.10E-04** | **4.17E-09** | -0.119 | 1.00E+00 | 5.26E-01 | 2.820 | 1.00E+00 | 1.10E-03 |
| HLA-DQB2 | 0.985 | 1.00E+00 | 1.19E-01 | 1.553 | 1.00E+00 | 6.49E-02 | 2.235 | 1.00E+00 | 5.69E-02 |
| HLA-DRA | **0.998** | **3.42E-06** | **1.30E-10** | NA | NA | NA | 1.826 | 1.00E+00 | 1.30E-04 |
| HLA-DRB1 | **1.133** | **6.71E-07** | **2.55E-11** | -0.104 | 1.00E+00 | 7.95E-01 | 1.718 | 1.00E+00 | 2.05E-04 |
| HLA-DRB5 | NA | NA | NA | -0.254 | 1.00E+00 | 5.29E-01 | 1.432 | 1.00E+00 | 1.12E-01 |
| HLA-E | 0.267 | 1.00E+00 | 4.50E-01 | 0.916 | 1.00E+00 | 1.20E-01 | 0.784 | 1.00E+00 | 2.58E-01 |
| HLA-F | 1.190 | 1.00E+00 | 3.42E-02 | 2.248 | 1.00E+00 | 6.33E-03 | 1.215 | 1.00E+00 | 9.41E-02 |
| HLA-G | NA | NA | NA | NA | NA | NA | NA | NA | NA |
| IL1A | NA | NA | NA | NA | NA | NA | NA | NA | NA |
| IL1B | NA | NA | NA | NA | NA | NA | NA | NA | NA |
| IL6 | NA | NA | NA | NA | NA | NA | -3.350 | 1.00E+00 | 2.49E-02 |
| IL10 | NA | NA | NA | NA | NA | NA | NA | NA | NA |
| IL12A | NA | NA | NA | NA | NA | NA | NA | NA | NA |
| IL12B | 4.031 | 1.00E+00 | 2.07E-03 | NA | NA | NA | 1.142 | 1.00E+00 | 4.78E-01 |
| IL15 | **2.525** | **3.50E-04** | **1.33E-08** | 0.691 | 1.00E+00 | 3.87E-02 | **1.475** | **5.94E-03** | **2.26E-07** |
| IL18 | 0.178 | 1.00E+00 | 9.17E-01 | NA | NA | NA | 1.174 | 1.00E+00 | 3.19E-01 |
| IL23A | NA | NA | NA | NA | NA | NA | NA | NA | NA |
| TNF | 0.694 | 1.00E+00 | 7.37E-01 | -0.932 | 1.00E+00 | 6.91E-01 | 0.458 | 1.00E+00 | 3.76E-01 |

**Extended Data Figure 1. Expression of top 30 cDC1-specific, top 30 cDC2-specific, and top 30 mregDC-specific genes in each cell type from snRNA-seq.** Genes are ordered from top to bottom by adjusted p-value (smallest to largest) and, if tied, by log2FC (largest to smallest) versus all cells (adjusted p-value <0.001) for cDC1 cells (A), cDC2 cells (B), and mregDC cells (C).

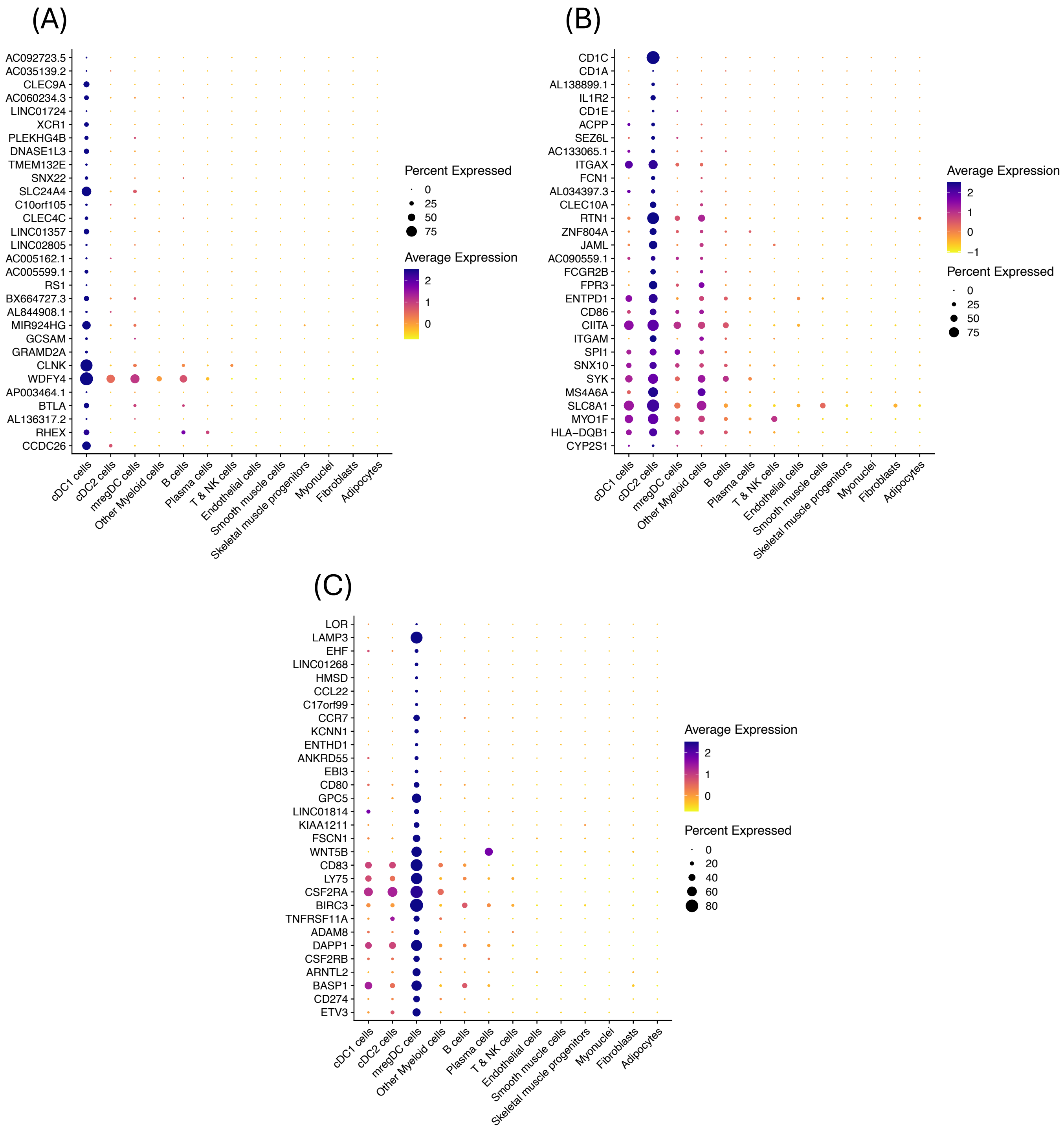

**Extended Data Figure 2. FeaturePlot of representative mDC gene expression in myeloid cells from snRNA-seq.** Showing expression of representative cDC1 genes (*CLEC9A*, *CLNK*, *DNASE1L3*, *XCR1*), cDC2 genes (*CD1C*, *CD1E*, *FCN1*, *IL1R2*), and mregDC genes (*ARNTL2*, *CCR7, FSCN1*, *LAMP3*).

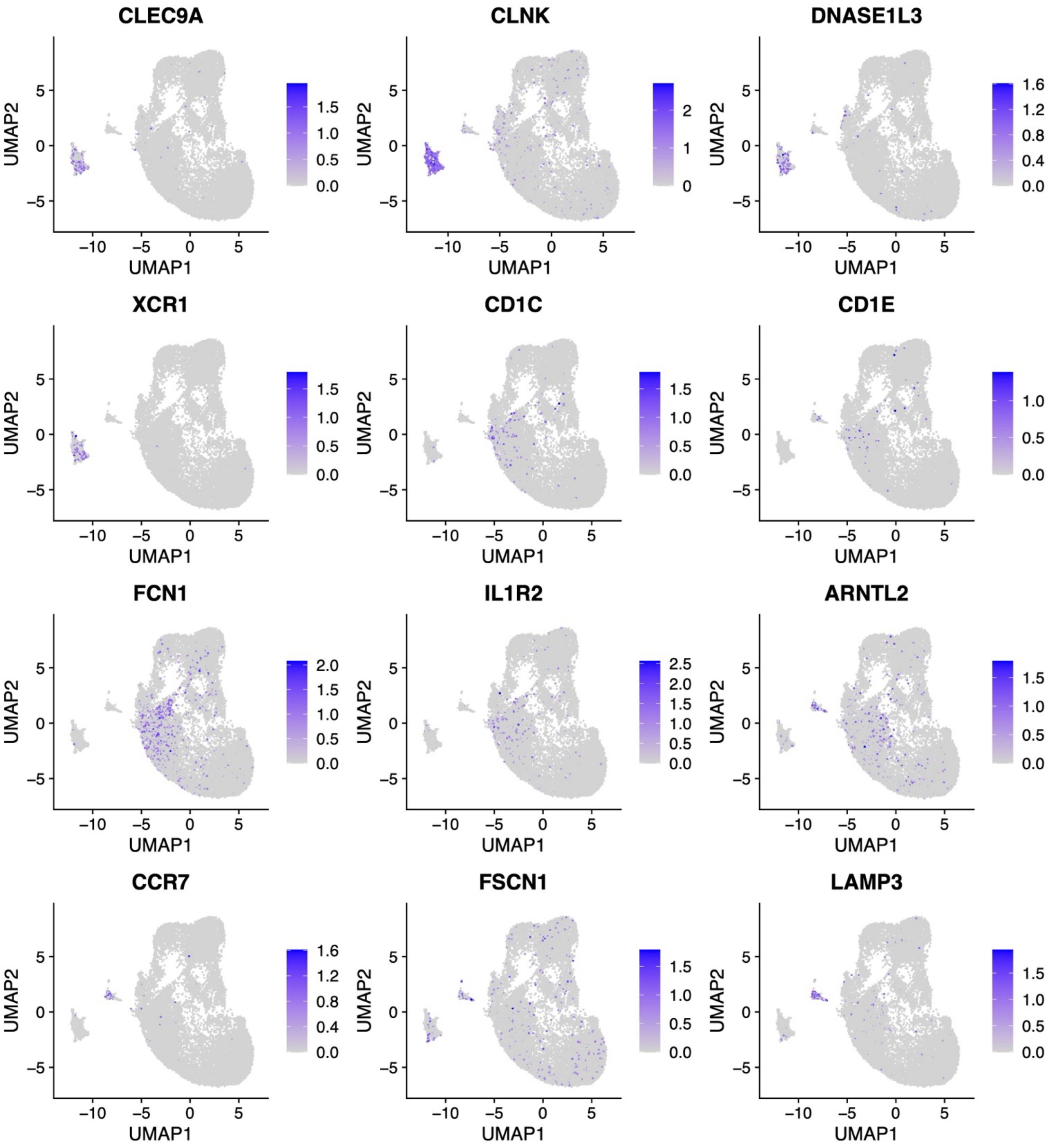

**Extended Data Figure 3. Correlation of mDC markers with each other in IBM samples from snRNA-seq.** Showing correlations of *CLEC9A* expression per myeloid cell with *CD1C* expression per myeloid cell (A), *CLEC9A* expression per myeloid cell with *LAMP3* expression per myeloid cell (B), and *CD1C* expression per myeloid cell with *LAMP3* expression per myeloid cell (C) among IBM samples.

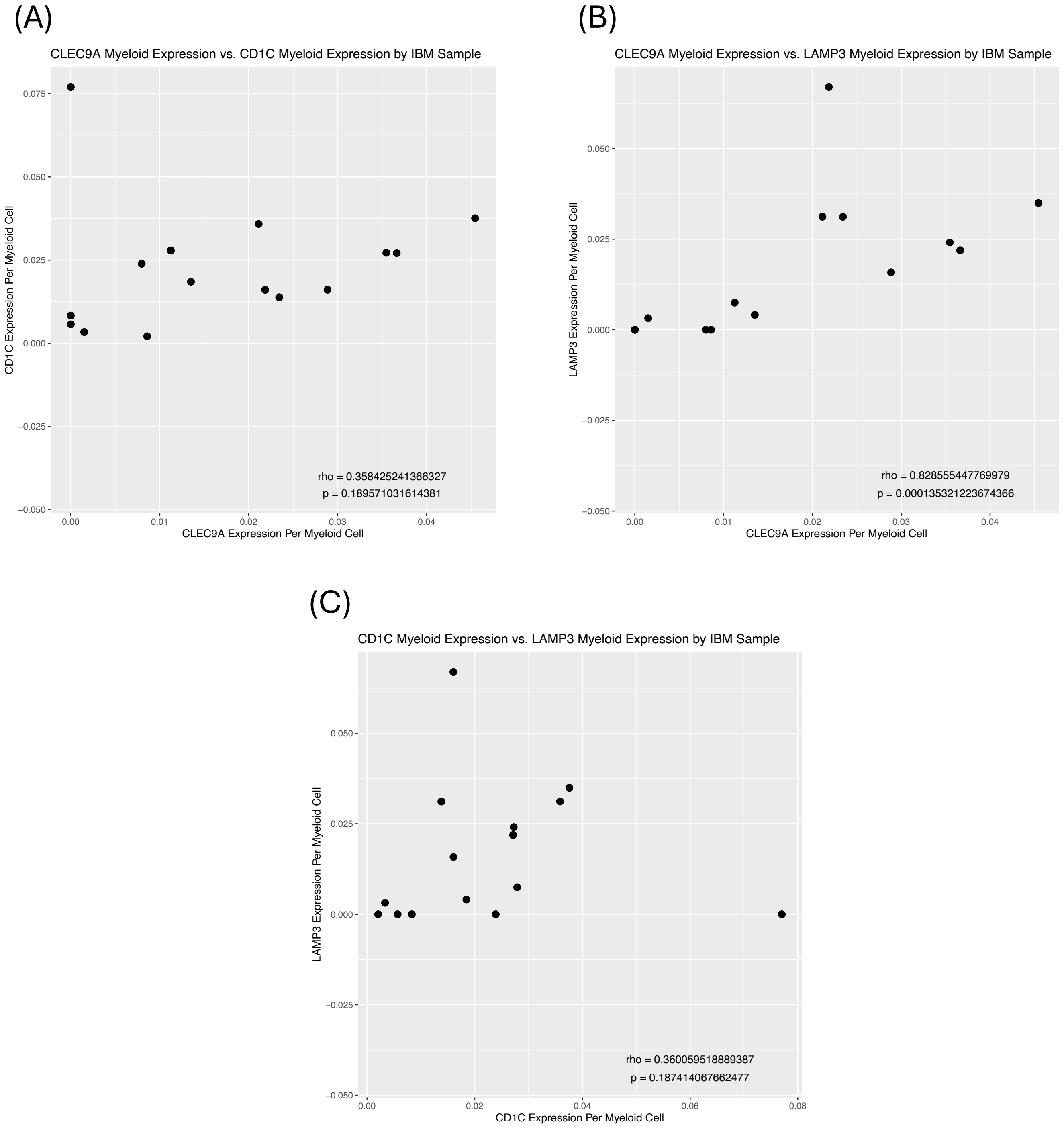

**Extended Data Figure 4. Expression of 58 inflammatory genes in each cell type from snRNA-seq.** Showing genes in IFN-II pathway, co-stimulatory genes, ligands of KLRG1 *(CDH1*, *CDH2),* co-inhibitory genes, HLA genes, and cytokines.

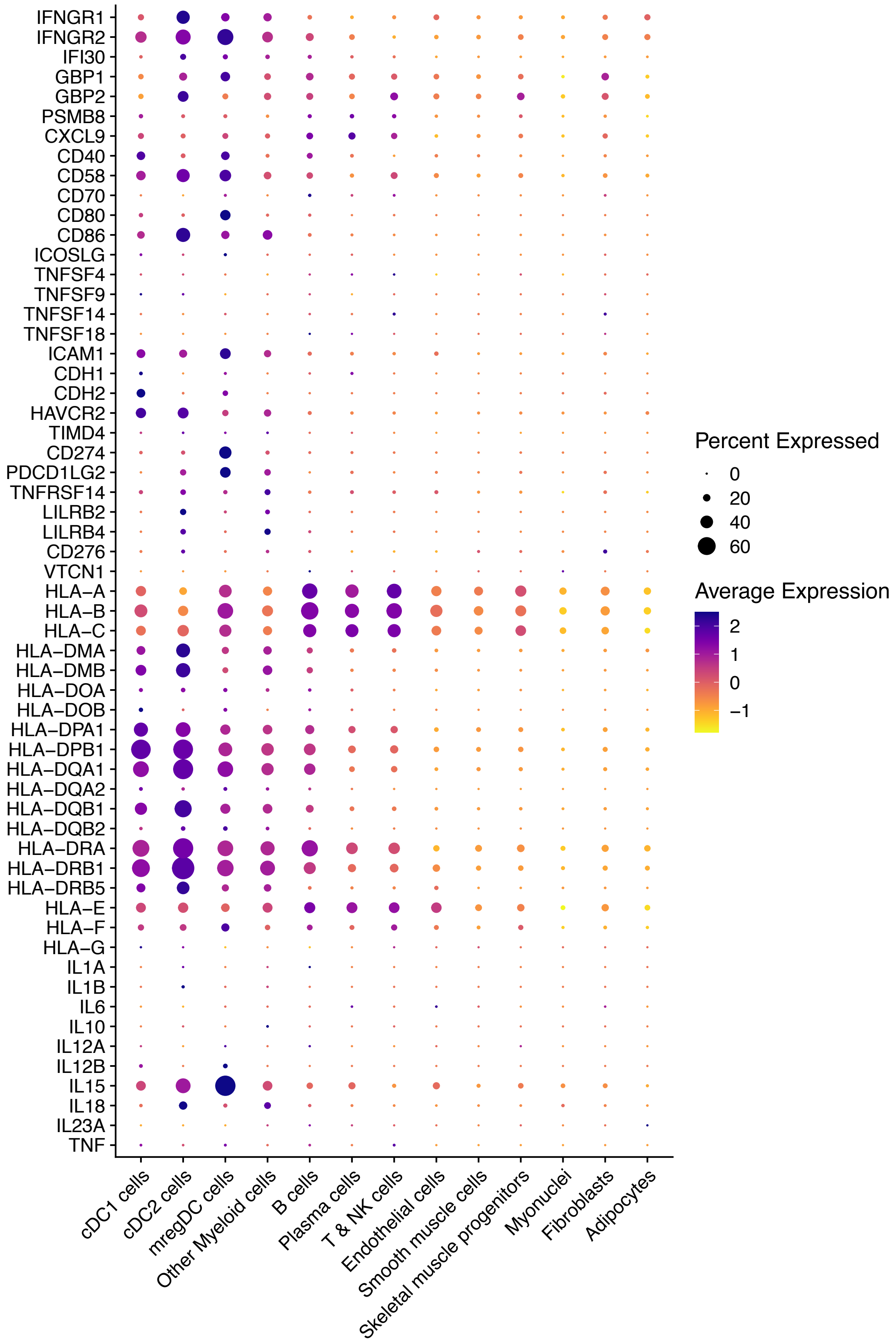

**Extended Data Figure 5. FeaturePlot of expression of *CDH1*, *CDH2*, and *IL12B* in myeloid cells from snRNA-seq.** Showing expression in myeloid cells from IBM samples (A) and non-IBM samples (B).

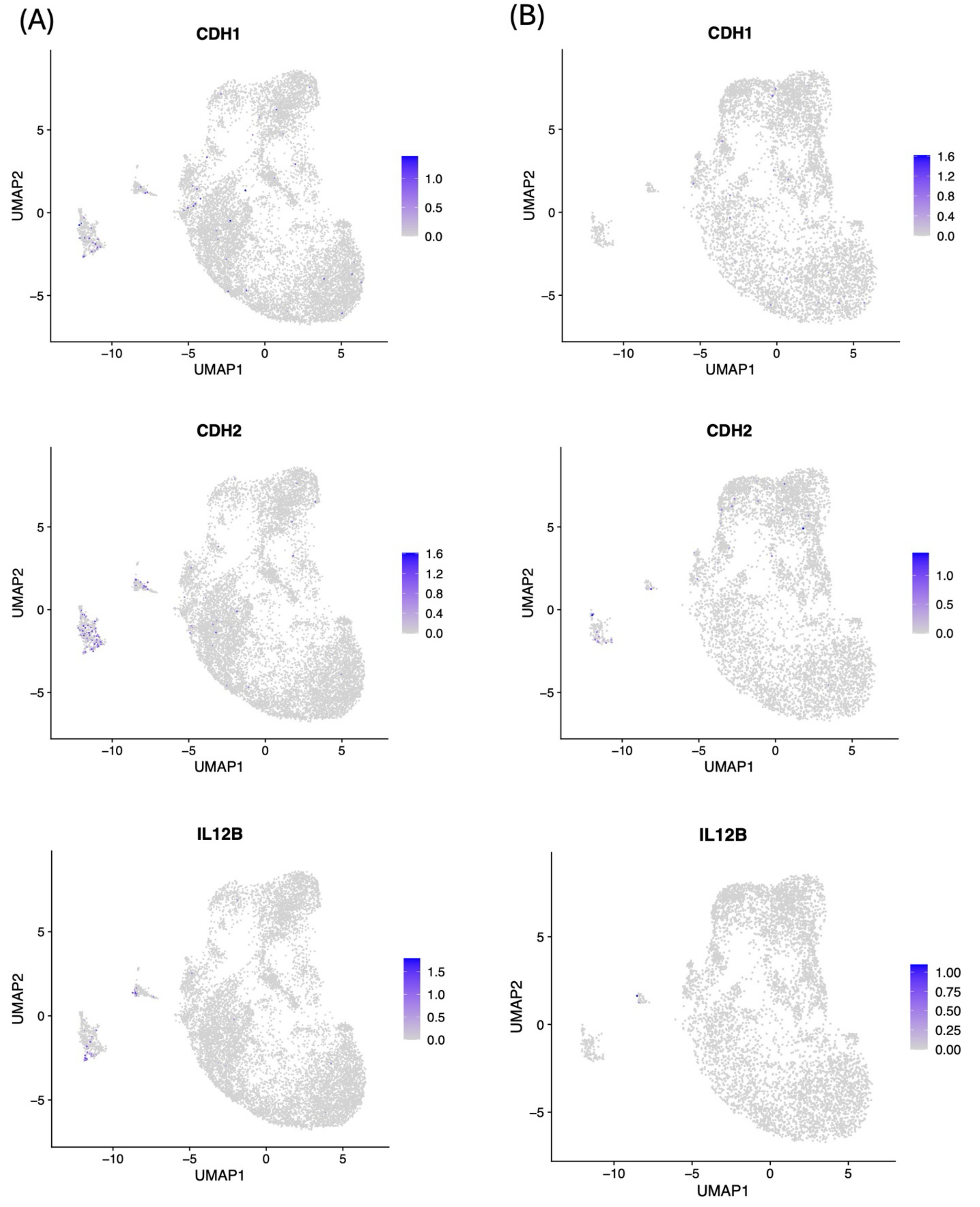

**Extended Data Figure 6. Correlation of mDC markers with Inflammatory T cell markers in all samples from snRNA-seq.** (A) Correlation of *KLRG1* expression per T/NK cell with *CLEC9A*, *CD1C*, and *LAMP3* expression per myeloid cell in all samples. (B) Correlation of *TBX21* expression per T/NK cell with *CLEC9A*, *CD1C*, and *LAMP3* expression per myeloid cell in all samples.

(A)

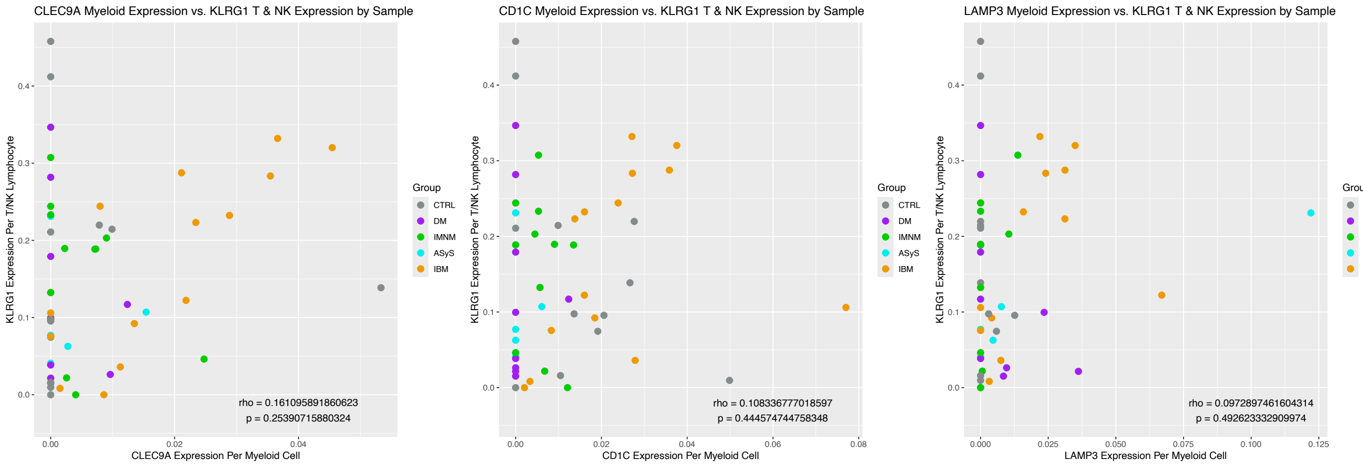


(B)

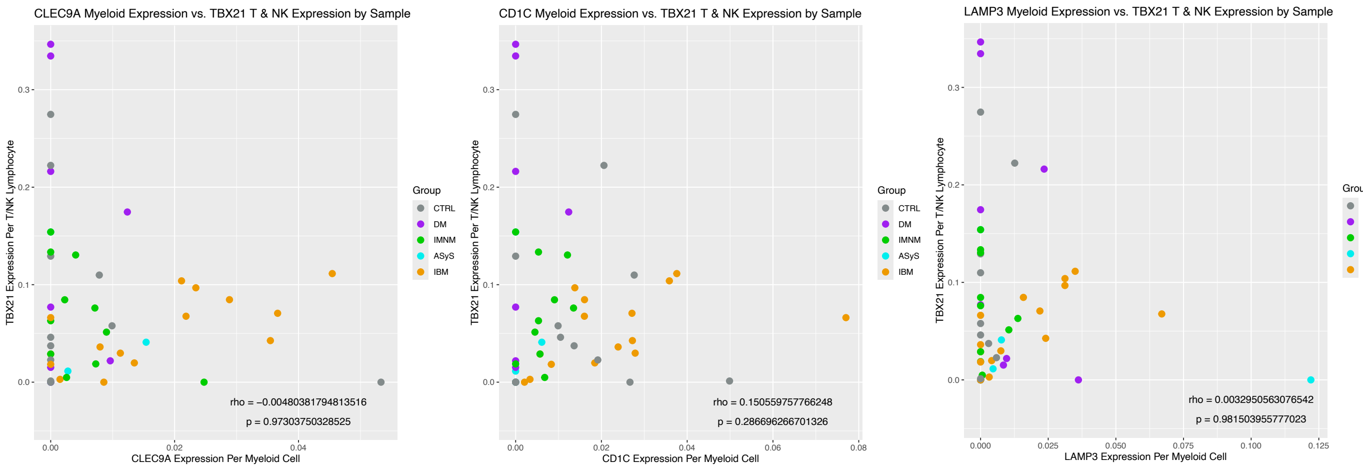

**Extended Data Figure 7. Correlation Heatmaps of specific mDC genes with each other from RNA-seq.** Correlation of expression of cDC1-specific genes (*CLEC9A*, *CLNK*, *DNASE1L3*, *XCR1*), cDC2 –specific genes (*CD1C*, *CD1E*, *FCN1, IL1R2*), and mregDC-specific genes (*ARNTL2*, *CCR7*, *FSCN1*, *LAMP3*) with each other. Correlations shown for IBM samples in dataset D (A), non-IBM myositis samples in dataset D (B), CTRL samples in dataset D (C), IBM samples in dataset E (D), and CTRL samples in dataset E (E).

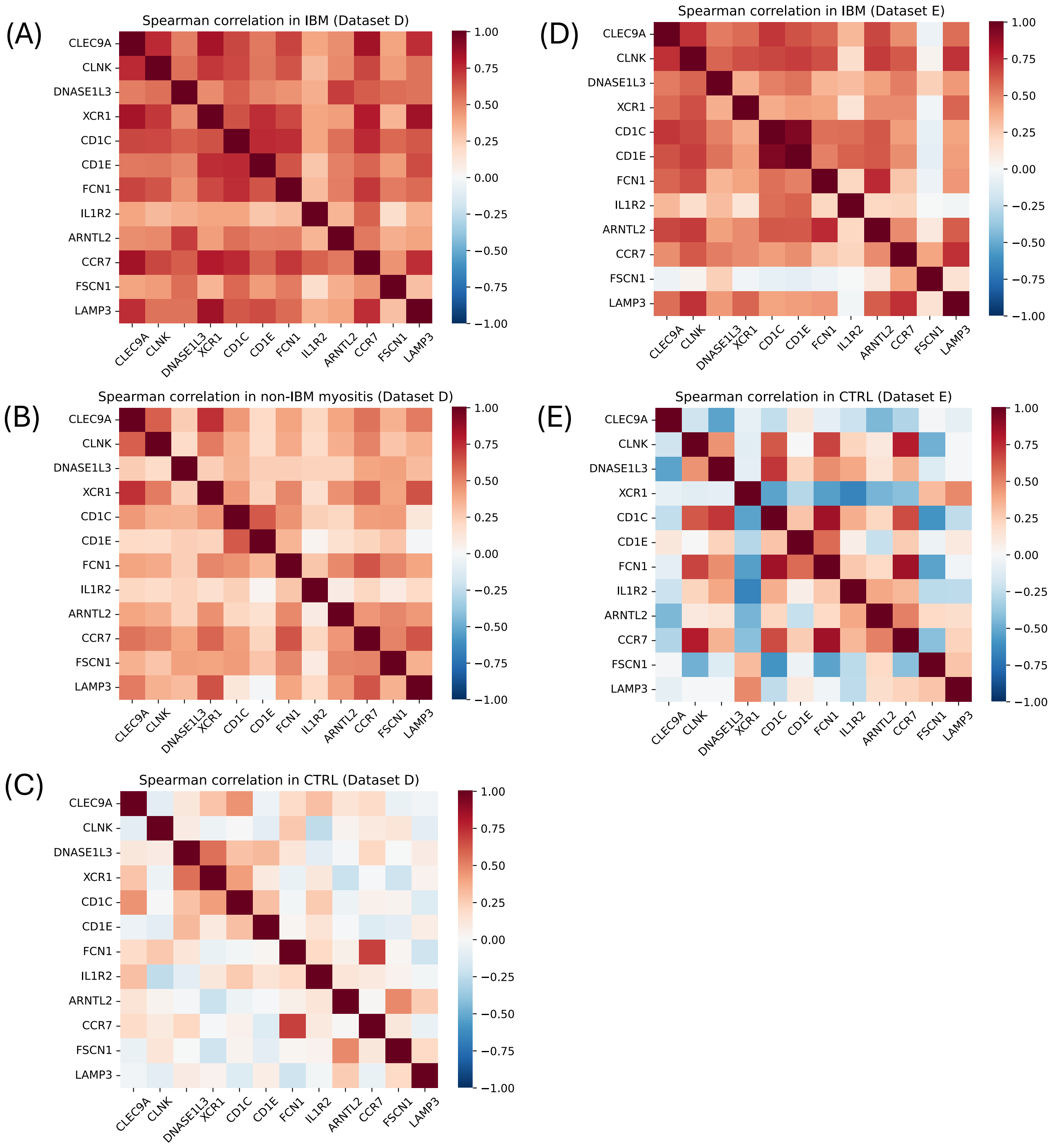

**Extended Data Figure 8. Differentially Correlated Genes in IBM for each specific mDC subset marker from RNA-seq.** Showing genes correlated with each mDC subset marker (*CLEC9A*, *CD1C*, *LAMP3*) in IBM patients from Dataset D with spearman correlation ρ >0.7 and Δρ >0.2 compared to that in non-IBM patients from Dataset D.

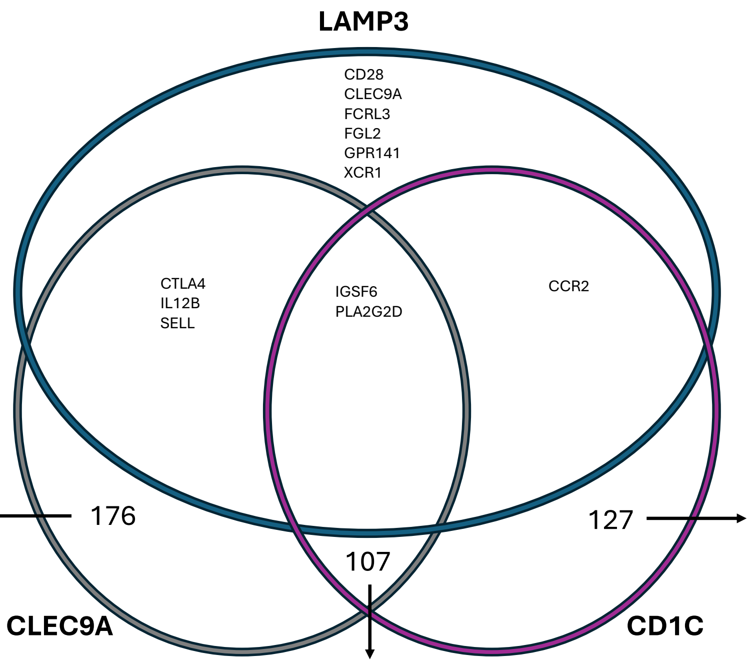


ACAP1
ADAM8
ADAP1
ADGRG5
AKNA
ANKRD22
ANO9
APOBEC3A
APOBEC3G
APOBEC3H
APOBR
APOL1
APOL2
APOL3
ARL4C
ASPHD2
BATF
BCL11B
BCL2L14
BICDL1
BTN2A1
BTN3A1
BTN3A2
BTN3A3
C11ORF21
C1QA
C1QB
C1QC
C4A
CASP1
CCDC88B
CCL17
CCL19
CCL2
CCL3
CCL4
CCL4L2
CCL8f
CD1D
CD244
CD3D
CD7
CD8A
CD8B
CFB
CLEC4E
CLNK
CORO1A
CST7
CTSW
CXCL10
CXCL11
CXCL13
CXCR2P1
CYP21A2
CYP2S1
CYSLTR2
DRAM1
ENSG00000266088.6
ENSG00000268088.1
ENSG00000272825.1
ENSG00000273272.2
EPSTI1
ETV7
EVL
FCGR3A
FCHO1
FGD2
FGD3
FMNL1-DT
FOXP3
FUT7
GALNT6
GBP1
GBP2
GBP4
GBP5
GBP6
GCH1
GJD3
GNA15
GPSM3
GZMB
GZMH
HK3
HLA-B
HLA-DQB2
HLA-DRB6
HLA-F
ICAM1
ICOS
IFI35
IFNG
IGFLR1
IKZF3
IL18BP
IL27RA
IRF1
IRF5
IRF9
JAK3
JAKMIP1
JAML
KCNN4
KLHDC7B
KLRK1
LAG3
LAMP3
LAT
LGALS9
LINC00539
LTB
MATK
MCOLN2
MIAT
MIR155HG
MMP25
MYO1G
NKG7
NLRC5
NOD2
PARP12
PARP14
PARVG
PATL2
PCED1B-AS1
PDCD1
PIK3R5
PIM2
PLA2G7
PLAAT4
PRF1
PSMB8
PSMB8-AS1
PSMB9
PSME1
PSME2
PSTPIP1
PTPRCAP
RAB37
RGL4
RUFY4
S1PR4
SCML4
SECTM1
SEMA7A
SH2D1A
SIGLEC1
SKAP1
SLA2
SLC15A3
SLFN12L
SNX20
SOWAHD
SP140L
SPOCK2
TAGAP
TAP1
TAP2
TBX21
THEMIS2
TMEM229B
TNFAIP2
TNFRSF14-AS1
TRAFD1
TRIM22
TTC16
TTYH3
TYMP
UBA7
UBD
UBE2L6
USP30-AS1
ZAP70
ZBED2
ZBP1

AIF1
ARHGAP30
ARHGAP9
BATF3
BIN2
BTN2A2
C15ORF48
CALHM6
CCL5
CCR1
CCR7
CD163L1
CD300LF
CD37
CD3E
CD52
CD6
CD74
CD96
CEACAM21
CIITA
CLEC10A
CLSTN3
COTL1
CTSZ
CXCL9
CXCR3
CXCR6
CYTH4
DEF6
DGKA
DOK2
FBP1
FCGR1A
FCGR1B
FCRL6
FERMT3
FMNL1
FPR3
FYB1
GBP1P1
GMIP
GPR132
GPR68
GRAP2
GZMA
HCK
HCST
HLA-DMB
HLA-DPA1
HLA-DPB1
HLA-DQA2
HLA-DRA
HLA-DRB1
IFI30
IKBKE
IL10RA
IL12RB1
IL2RB
IL2RG
IL4I1
ITGAX
ITGB2
ITGB2-AS1
LAPTM5
LCP1
LGALS2
LILRB1
LILRB4
LINC01871
LST1
LYZ
MMP25-AS1
MMP9
MYO1F
NCF1
NCKAP1L
PIK3CD
PLCB2
PLEK
PRKCB
PTAFR
PTPN6
PTPRE
RAC2
RASAL3
RPS6KA1
RUNX3
SAMD3
SASH3
SELPLG
SERPINA1
SH2D2A
SH3BP1
SLAMF8
SPI1
STXBP2
TCF7
TMC8
TNF
TNFSF14
TRAC
TRAF1
TRBC2
UBASH3A
WAS
ZNF683

ADA2
ADAP2
ADCY7
AOAH
APOBEC3C
APOL4
ARHGAP15
ARHGAP25
ARRB2
BTK
C16ORF54
C2
CASP4
CCL13
CCR5
CD1A
CD1B
CD2
CD33
CD3G
CD4
CD48
CD5
CD53
CD83
CHST11
CRTAM
CSF1
CTSS
CXCL16
DENND1C
DOCK10
DOCK2
DOK3
DPEP2
EFHD2
ENSG00000255197.7
ENSG00000259834.1
ENSG00000260997.1
EVI2B
FASLG
FCN1
GLIPR2
GPR84
HACD4
HLA-DMA
HLA-DOA
HLA-DQA1
HLA-DQB1
HPSE
IKZF1
IL21R
INPP5D
IRF8
ITGAL
ITGAM
ITGB7
KLRB1
KLRG1
LAT2
LCK
LCP2
LILRB2
LILRB3
LINC00892
LPAR5
LPXN
LYN
MIR3142HG
MPEG1
MS4A6A
MXRA5
NCF2
NCR3
NFKBIE
NLRC3
NRROS
PENK
PIK3CG
PIK3R6
PLAU
PPM1M
PPP1R18
PRR5L
PSD4
PTPN22
PTPN7
PTPRC
RASSF2
RBP5
RGS19
RIN3
RNASET2
RTN1
SAMSN1
SEPTIN6
SH3BGRL3
SIGLEC7
SLAMF7
SLC29A3
SP140
SPINT1
SPN
STK10
SUSD3
TBXAS1
TFEC
TGFB1
TIFAB
TNFAIP8L2
TNFSF13B
TOX
TRAF3IP3
TRAT1
TRBC1
TREM2
TRG-AS1
TRGC1
TRGC2
TRPV2
TSPOAP1-AS1
TVP23A
TYROBP
VAV1
WDFY4
WIPF1
XCL2

**Extended Data Figure 9. Median Expression and Correlation Heatmaps of specific mDC genes in PM-CD8 and PM-Mito patients from RNA-seq.** (A) Median Expression of mDC-specific genes for groups included in RNA-seq datasets D and E, as well as PM-CD8 and PM-Mito patients. (B/C) Correlation of expression of cDC1-specific genes (*CLEC9A*, *CLNK*, *DNASE1L3*, *XCR1*), cDC2 –specific genes (*CD1C*, *CD1E*, *FCN1*, *IL1R2*), and mregDC-specific genes (*ARNTL2*, *CCR7*, *FSCN1*, *LAMP3*) versus markers of muscle regeneration, mature muscle, the IFN-II pathway, and immune cells in PM-CD8 patients (B) and PM-Mito patients (C).

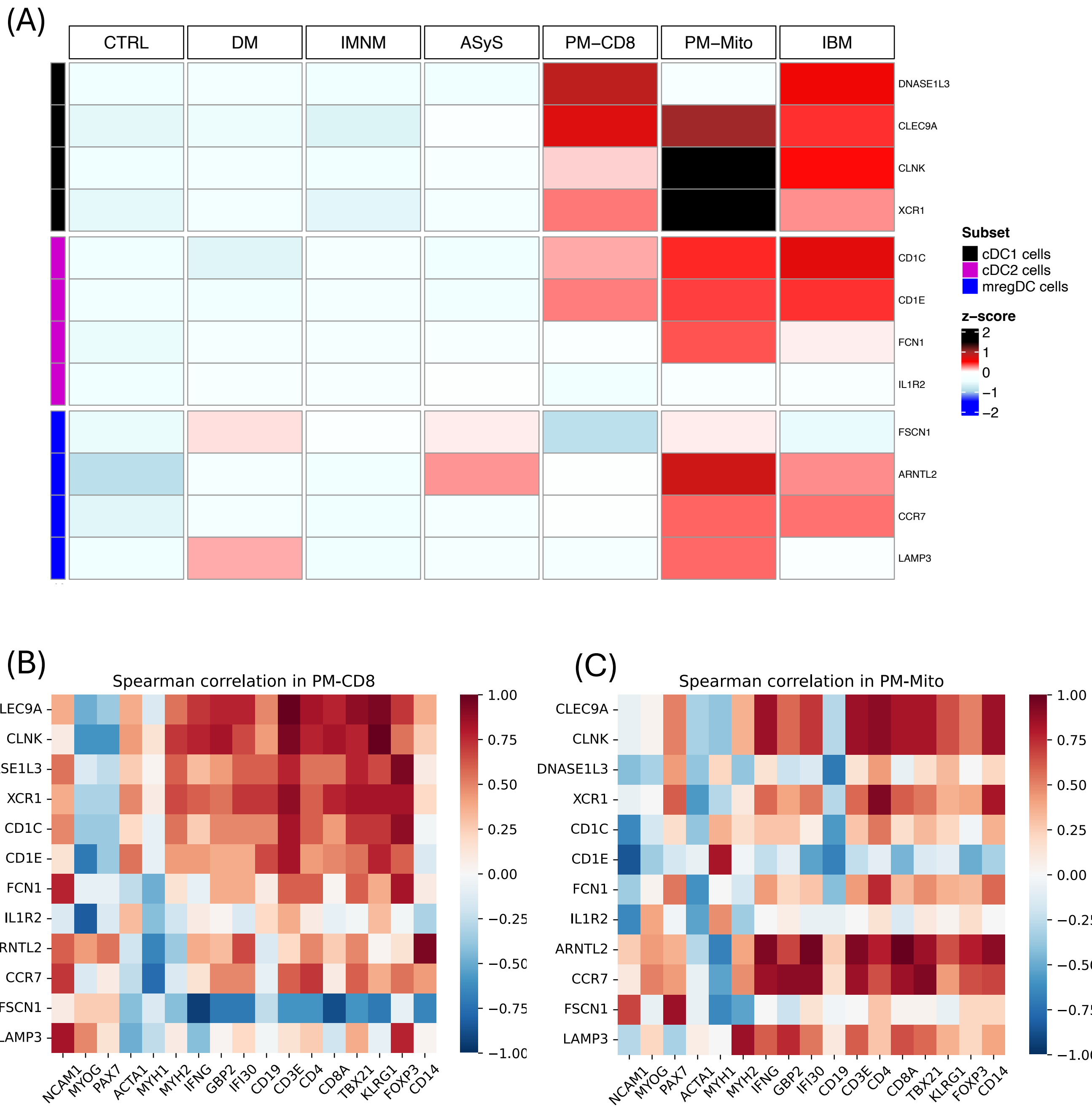
