## Supplementary Figures for "Activated Dendritic Cell Subsets Characterize Muscle of Inclusion Body Myositis Patients and Correlate with KLRG1+ and TBX21+ CD8+ T cells"

**Supplementary Figure 1. UMAP clustering of all filtered cells from snRNA-seq.** UMAP representations of original cell clusters generated by FindClusters function (A), and cells labeled by dataset they originated from (B).

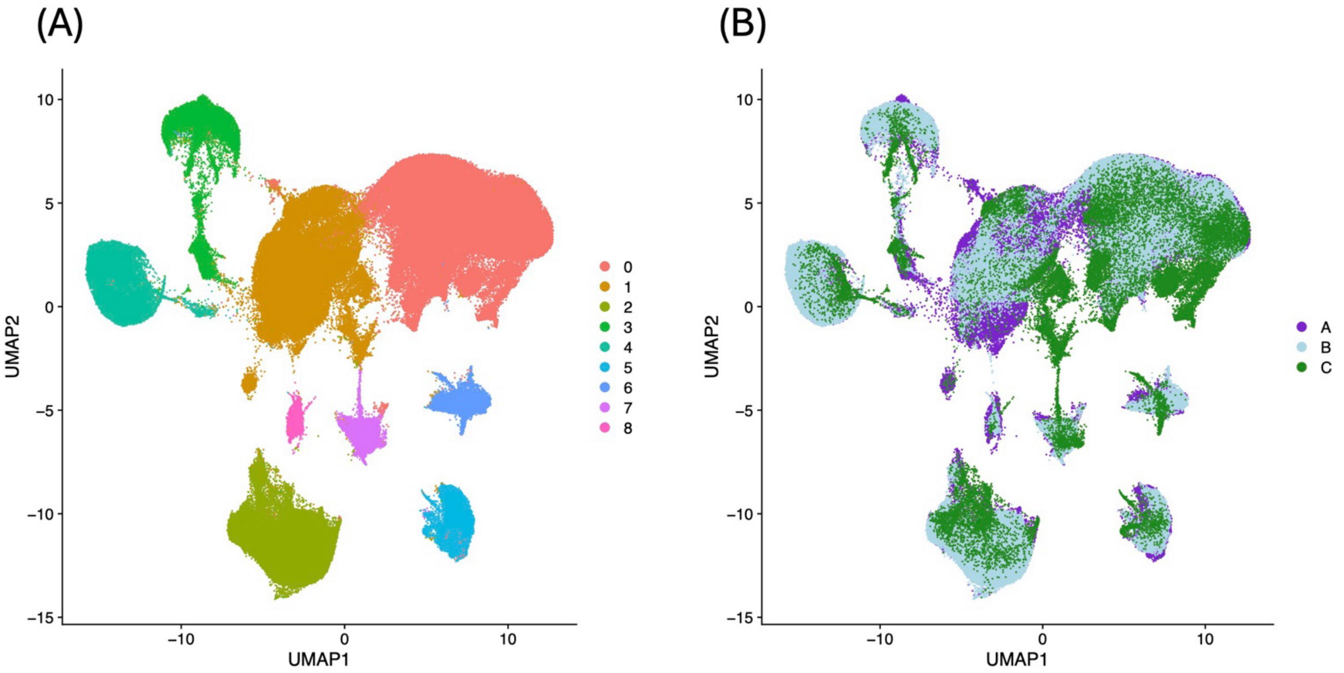

**Supplementary Figure 2. FeaturePlot of representative genes for each cell cluster from snRNA-seq.**

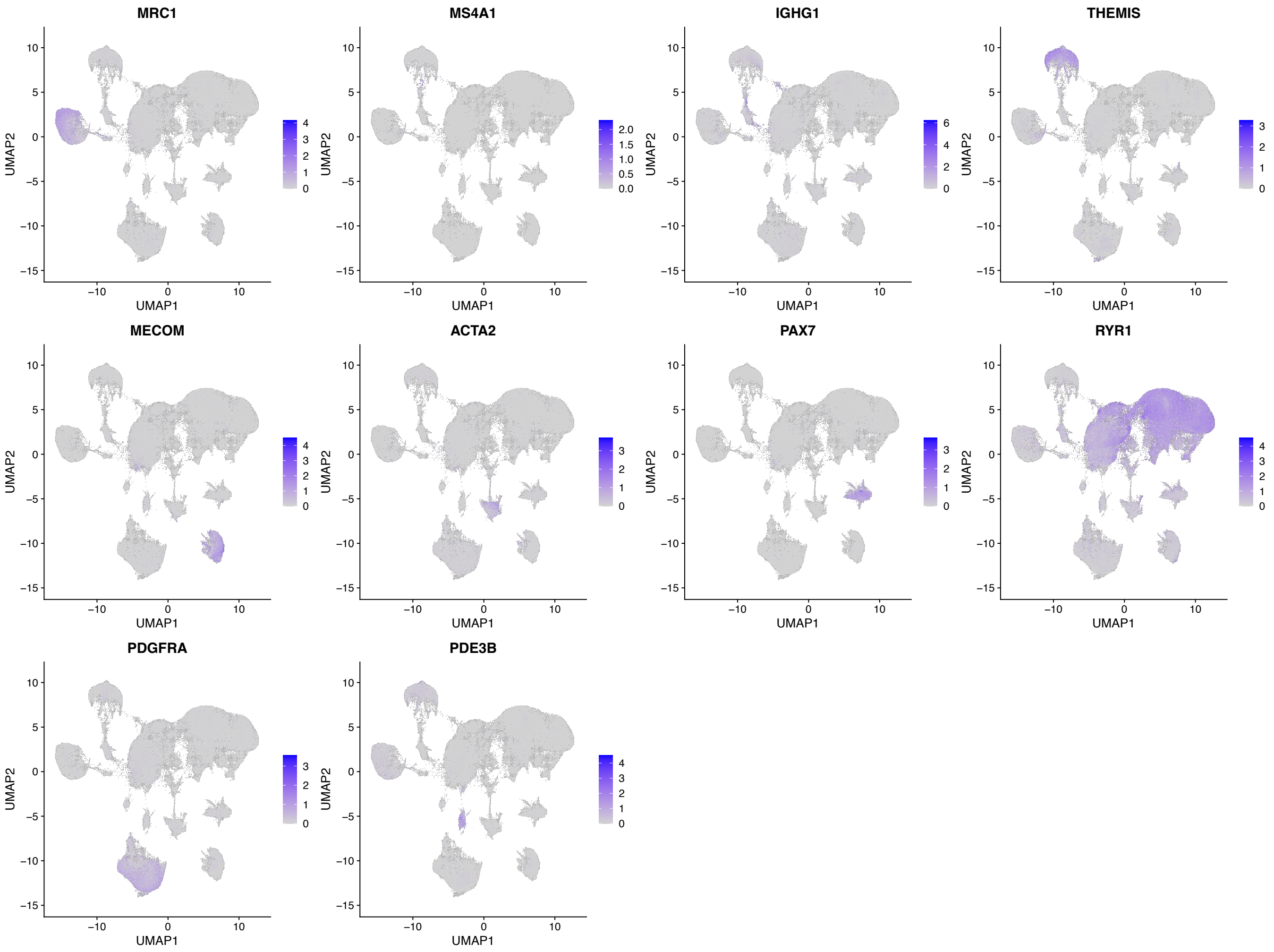

**Supplementary Figure 3. UMAP clustering of Lymphocytes from snRNA-seq.** UMAP representations of original cell clusters generated by FindClusters function (A), and corresponding named clusters (B).

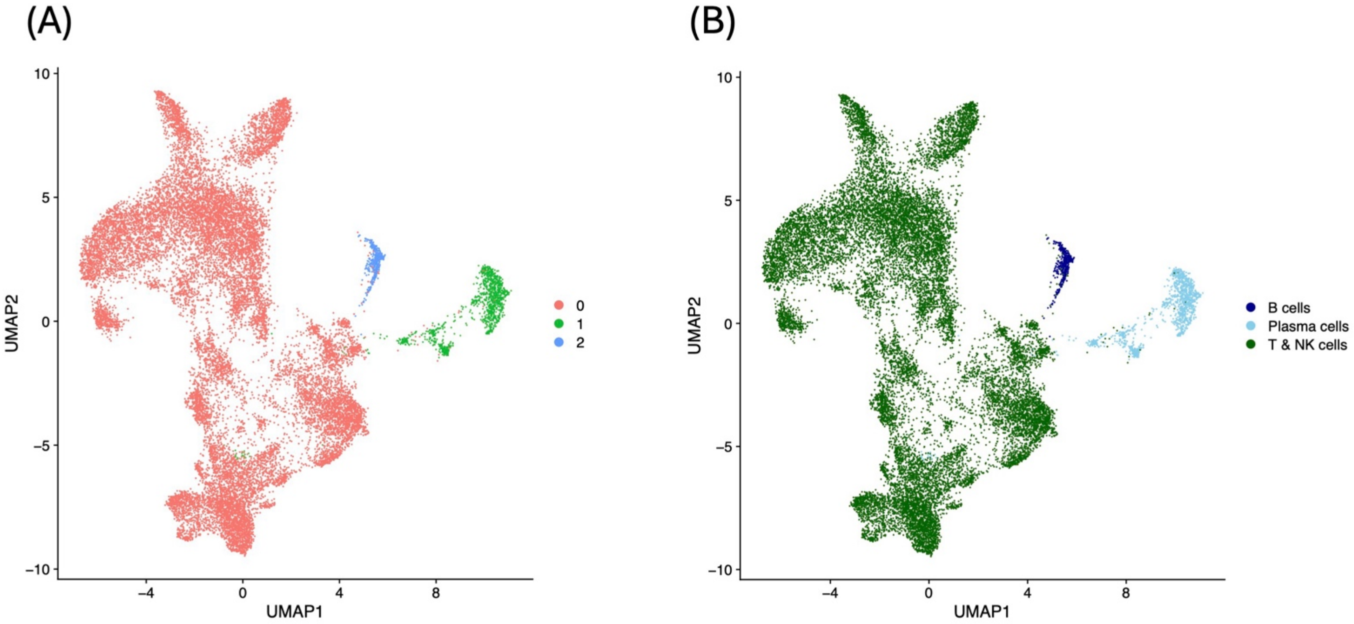

**Supplementary Figure 4. FeaturePlot of lymphocyte gene expression for lymphocyte clusters from snRNA-seq.** Showing expression of T cell markers (*THEMIS*, *ITGAE*, *CD3D*, *CD4*, *CD8A*, *FOXP3*), NK cell marker (*NCR1*), B cell markers (*MS4A1*, *PAX5*), and Plasma cell markers (*IGHG1*, *IGKC*, *PRDM1*).

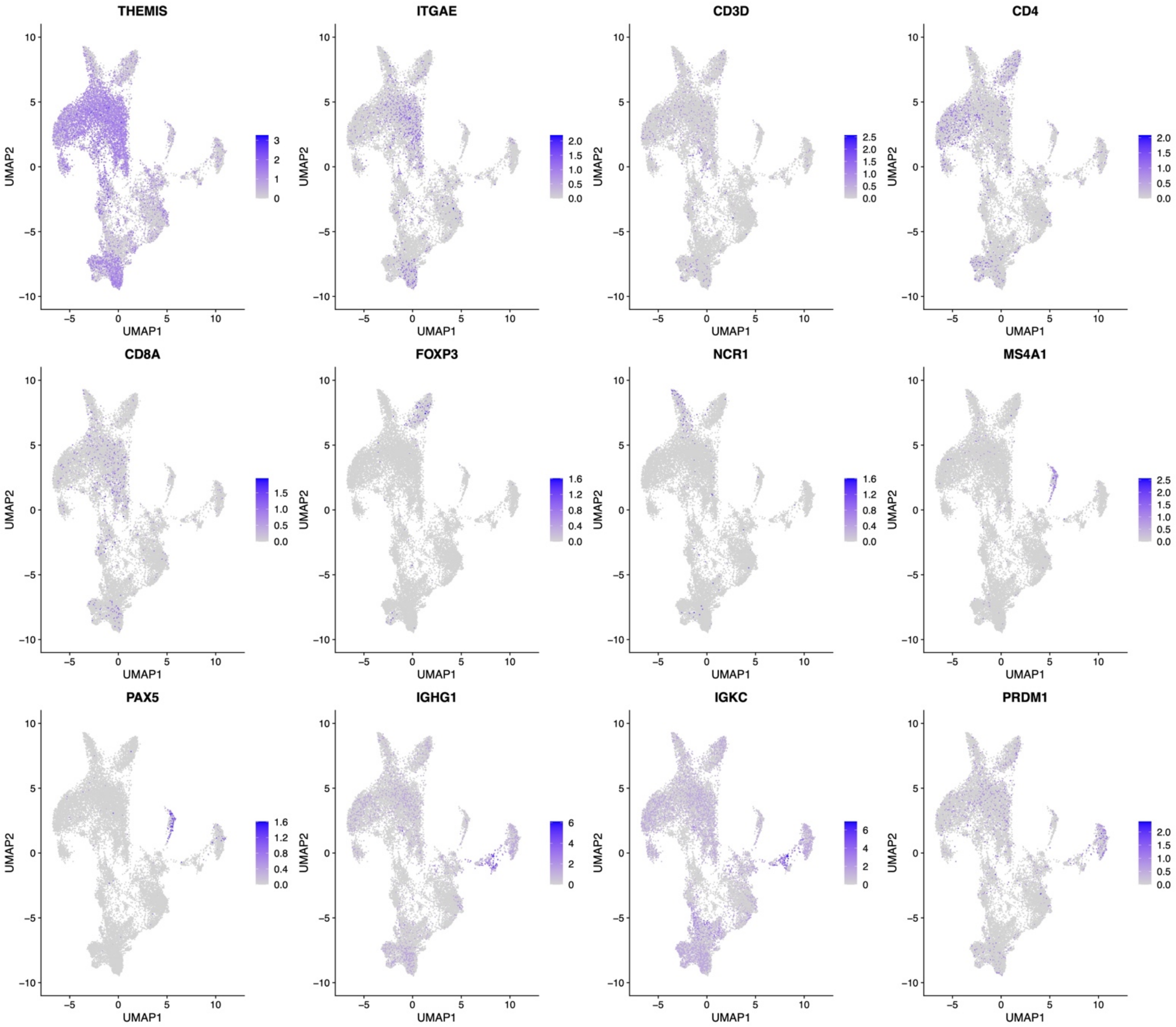

**Supplementary Figure 5. UMAP clustering of Myeloid cells from snRNA-seq.** UMAP representations of original cell clusters generated by FindClusters function (A), cells labeled by group (B), and cells labeled by dataset they originated from (C).

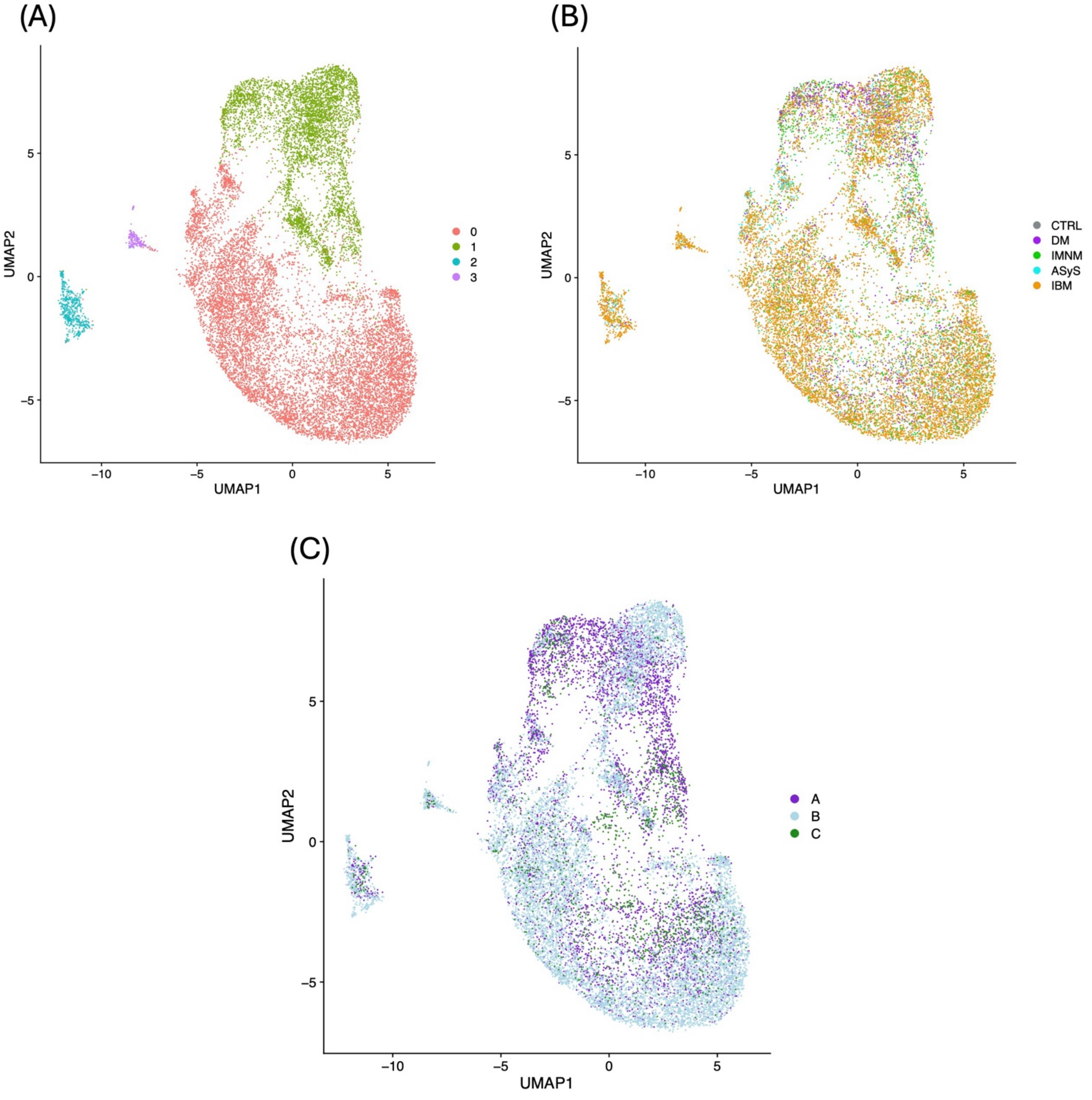
